# Global biobank analyses provide lessons for developing polygenic risk scores across diverse cohorts

**DOI:** 10.1101/2021.11.18.21266545

**Authors:** Ying Wang, Shinichi Namba, Esteban Lopera, Sini Kerminen, Kristin Tsuo, Kristi Läll, Masahiro Kanai, Wei Zhou, Kuan-Han Wu, Marie-Julie Favé, Laxmi Bhatta, Philip Awadalla, Ben Brumpton, Patrick Deelen, Kristian Hveem, Valeria Lo Faro, Reedik Mägi, Yoshinori Murakami, Serena Sanna, Jordan W. Smoller, Jasmina Uzunovic, Brooke N. Wolford, Global Biobank Meta-analysis Initiative, Cristen Willer, Eric R. Gamazon, Nancy J. Cox, Ida Surakka, Yukinori Okada, Alicia R. Martin, Jibril Hirbo

**Affiliations:** Analytic and Translational Genetics Unit, Massachusetts General Hospital, Boston, MA 02114, USA; Stanley Center for Psychiatric Research, Broad Institute of MIT and Harvard, Cambridge, MA 02142, USA; Program in Medical and Population Genetics, Broad Institute of MIT and Harvard, Cambridge, MA 02142, USA; Department of Statistical Genetics, Osaka University Graduate School of Medicine, Suita 565-0871, Japan; University of Groningen, UMCG, Department of Genetics, Groningen, the Netherlands; Institute for Molecular Medicine Finland, FIMM, HiLIFE, University of Helsinki, Helsinki, Finland; Estonian Genome Centre, Institute of Genomics, University of Tartu, Tartu, Estonia; Department of Biomedical Informatics, Harvard Medical School, Boston, MA, USA; Department of Statistical Genetics, Osaka University Graduate School of Medicine, Suita, Japan; Department of Computational Medicine and Bioinformatics, University of Michigan, Ann Arbor MI, 48103, USA; Ontario Institute for Cancer Research, Toronto, Ontario, Canada; K.G. Jebsen Center for Genetic Epidemiology, Department of Public Health and Nursing, NTNU, Norwegian University of Science and Technology, Trondheim, 7030, Norway; Department of Molecular Genetics, University of Toronto, Toronto, ON, Canada; HUNT Research Centre, Department of Public Health and Nursing, NTNU, Norwegian University of Science and Technology, Levanger, 7600, Norway; Clinic of Medicine, St. Olavs Hospital, Trondheim University Hospital, Trondheim, 7030, Norway; Oncode Institute, Utrecht, The Netherlands; Department of Ophthalmology, University Medical Center Groningen, University of Groningen, Groningen, The Netherlands; Department of Clinical Genetics, Amsterdam University Medical Center (AMC), Amsterdam, The Netherlands; Department of Immunology, Genetics and Pathology, Science for Life Laboratory, Uppsala University, Uppsala, Sweden; Division of Molecular Pathology, Institute of Medical Science, The University of Tokyo, Tokyo, Japan; Institute for Genetics and Biomedical Research (IRGB), National Research Council (CNR), Cagliari 09100, Italy; Psychiatric and Neurodevelopmental Genetics Unit, Center for Genomic Medicine, Massachusetts General Hospital, Boston, MA 02114, USA; Department of Internal Medicine, University of Michigan, Ann Arbor, MI 48109, USA; Department of Biostatistics and Center for Statistical Genetics, University of Michigan, Ann Arbor, MI 48109, USA; Department of Human Genetics, University of Michigan, Ann Arbor, MI 48109, USA; Department of Medicine, Division of Genetic Medicine, Vanderbilt University School of Medicine, Nashville, TN, USA; MRC Epidemiology Unit, University of Cambridge, Cambridge, UK; Vanderbilt Genetics Institute, Vanderbilt University Medical Center, Nashville, TN, USA; Laboratory for Systems Genetics, RIKEN Center for Integrative Medical Sciences, Yokohama, Japan; Laboratory of Statistical Immunology, Immunology Frontier Research Center (WPI-IFReC), Osaka University, Suita 565-0871, Japan; Department of Genome Informatics, Graduate School of Medicine, the University of Tokyo, Tokyo 113-0033, Japan; Center for Infectious Disease Education and Research (CiDER), Osaka University, Suita 565-0871, Japan

**Keywords:** Global-biobank meta-analysis initiative (GBMI), polygenic risk scores (PRS), multi-ancestry genetic prediction, accuracy heterogeneity

## Abstract

With the increasing availability of biobank-scale datasets that incorporate both genomic data and electronic health records, many associations between genetic variants and phenotypes of interest have been discovered. Polygenic risk scores (PRS), which are being widely explored in precision medicine, use the results of association studies to predict the genetic component of disease risk by accumulating risk alleles weighted by their effect sizes. However, few studies have thoroughly investigated best practices for PRS in global populations across different diseases. In this study, we utilize data from the Global-Biobank Meta-analysis Initiative (GBMI), which consists of individuals from diverse ancestries and across continents, to explore methodological considerations and PRS prediction performance in 9 different biobanks for 14 disease endpoints. Specifically, we constructed PRS using heuristic (pruning and thresholding, P+T) and Bayesian (PRS-CS) methods. We found that the genetic architecture, such as SNP-based heritability and polygenicity, varied greatly among endpoints. For both PRS construction methods, using a European ancestry LD reference panel resulted in comparable or higher prediction accuracy compared to several other non-European based panels; this is largely attributable to European descent populations still comprising the majority of GBMI participants. PRS-CS overall outperformed the classic P+T method, especially for endpoints with higher SNP-based heritability. For example, substantial improvements are observed in East-Asian ancestry (EAS) using PRS- CS compared to P+T for heart failure (HF) and chronic obstructive pulmonary disease (COPD). Notably, prediction accuracy is heterogeneous across endpoints, biobanks, and ancestries, especially for asthma which has known variation in disease prevalence across global populations. Overall, we provide lessons for PRS construction, evaluation, and interpretation using the GBMI and highlight the importance of best practices for PRS in the biobank-scale genomics era.

## Introduction

Population- and hospital-based biobanks are increasingly coupling genomic and electronic health record data at sufficient scale to evaluate the potential of personalized medicine^1^. The growth of these paired datasets enables genome-wide association studies (GWAS) to estimate increasingly precise genetic effect sizes contributing to disease risk. In turn, GWAS summary statistics can be used to aggregate the effects of many genetic markers (usually in the form of single -nucleotide polymorphisms, SNPs) to estimate individuals’ genetic predispositions for complex diseases via polygenic risk scores (PRS). As GWAS power has increased, PRS accuracy has also improved, with PRS for some traits having comparable accuracies to independent biomarkers already routinely used in clinical risk models^2^. Consequently, several areas of medicine have already begun investigating the potential for integrating PRS alongside other biomarkers and information currently used in clinical risk models^3–5^. However, evidence of clinical utility for PRS across disease areas is currently limited or inconsistent^2,6–8^. Furthermore, many methods have been developed to compute PRS, each with different strengths and weaknesses^9–11^. Thus, guidelines that delineate best practices while considering a range of real-world healthcare settings and disease areas are critically needed.

Best practices for PRS are critical but lacking for a range of considerations that have been shown to contribute to variability in accuracy and interpretation. These include guidance for variable phenotype definitions and precision for both discovery GWAS and target populations, which varies with cohort ascertainment strategy, geography, environmental exposures and other common covariates^12–14^. Other considerations include varying genetic architectures, statistical power of the discovery GWAS, and PRS methods, which vary in which variants (generally in the form of SNPs) are included and how weights are calculated^9,15^. A particularly pernicious issue requiring best practices is regarding maximizing generalizability of PRS accuracy among ancestry groups^16,17^. Developing best practices for PRS therefore requires harmonized genetic data spanning diverse phenotypes, participants, and ascertainment strategies.

To facilitate the development of best practices, we evaluate several considerations for PRS in the Global Biobank Meta-analysis Initiative (GBMI). GBMI brings together population- and hospital- based biobanks developed in twelve countries spanning four different continents: North America (USA, Canada), East Asia (Japan and China), Europe (Iceland, UK, Estonian, Finland, Scotland, Norway and Netherlands) and Oceania (Australia). GBMI aggregates paired genetic and phenotypic data from >2.1 million individuals across diverse ancestries, including: ∼1.4 million Europeans (EUR), ∼18,000 Admixed Americans (AMR), ∼1,600 Middle Eastern (MID), ∼31,000 Central and South Asians (CSA), ∼341,000 East Asians (EAS) and ∼33,000 Africans (AFR). Biobanks have collated phenotype information through different sources including electronic health records, self-report data from epidemiological survey questionnaires, billing codes, doctors’ narrative notes, and death registries. Detailed description of each biobank is found in Zhou et al.^18^.

Here we outline a framework for PRS analyses of multi-ancestry GWAS across multiple biobanks, as shown in **Figure 1**. The endpoints examined are: asthma, chronic obstructive pulmonary disease (COPD), heart failure (HF), stroke, acute appendicitis (AcApp), venous thromboembolism (VTE), gout, appendectomy, primary open-angle glaucoma (POAG), uterine cancer (UtC), abdominal aortic aneurysm (AAA), idiopathic pulmonary fibrosis (IPF), thyroid cancer (ThC) and hypertrophic or obstructive cardiomyopathy (HCM), for which the phenotype definitions can be found in Zhou et al.^18^. Those 14 endpoints represent the pilot effort of GBMI, which greatly vary in disease prevalence. It ranges from <1% for AAA, IPF, ThC and HCM to ∼6% for COPD and ∼9% for asthma. Some endpoints (for example, appendectomy which can be extracted from EHR procedure codes) have not been broadly studied in previous GWAS studies. By evaluating PRS across 14 endpoints (**Table S1 and Table S2**) and 9 biobanks, we review and explore practical considerations for three steps: genetic architecture estimation, PRS method optimization and selection, and evaluation of PRS accuracy. Our framework applies to biobank-scale resources with both homogenous and diverse ancestries.

**Figure 1.**
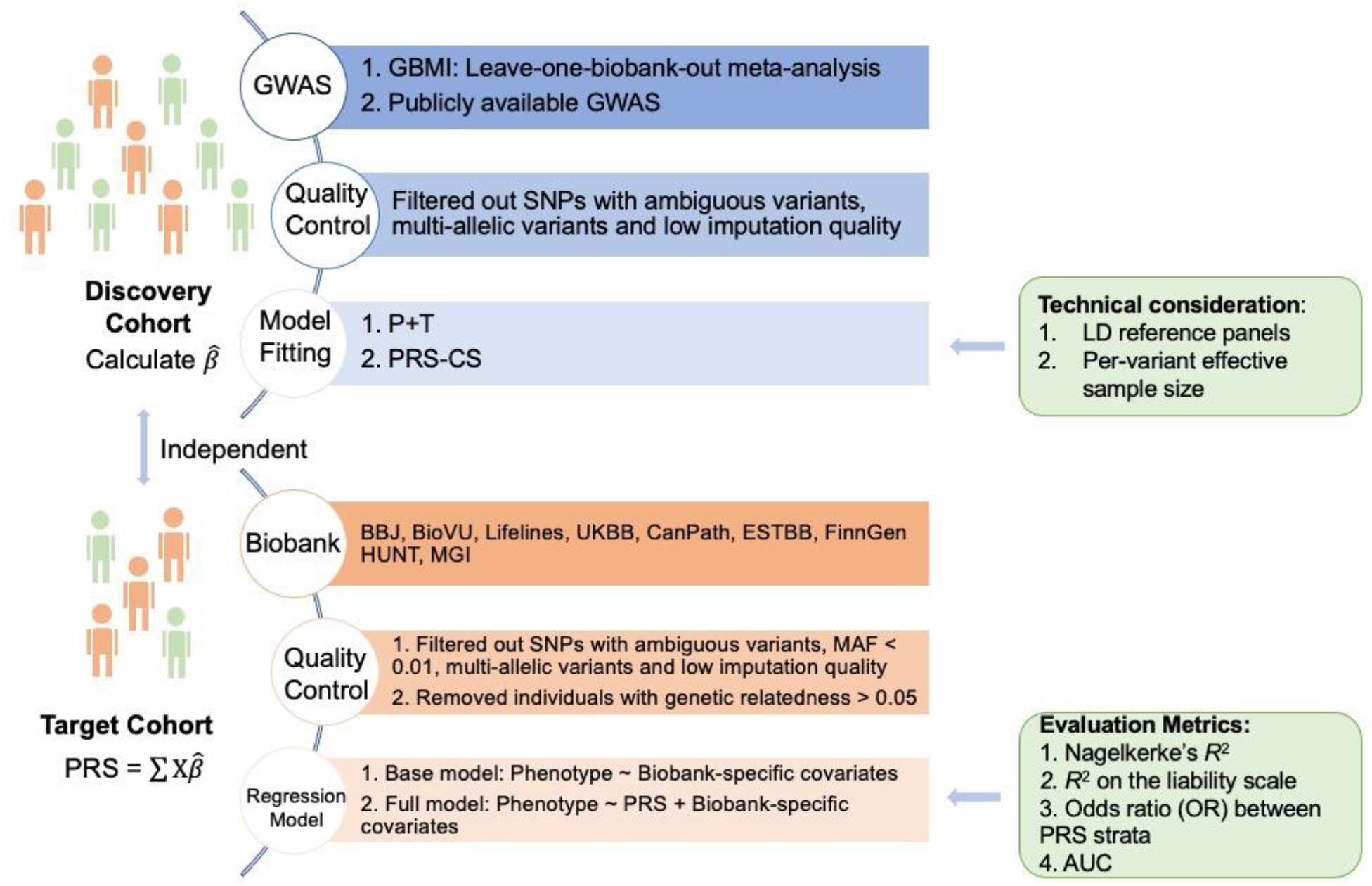
Overview of the study framework.

## Results

The diverse ancestries included in GBMI accounted for different proportions ranging from ∼76.4% for EUR, 0.1% for MID, 1.0% for AMR, 1.7% for CSA, 18.9% for EAS and 1.8% for AFR. We explored the genetic architecture of 14 endpoints using GWAS summary statistics from all ancestries and EUR only in GBMI^19^. We used leave-one-biobank-out meta-analyzed GWAS in GBMI as our primary discovery datasets for the following PRS analyses. The ancestry compositions of discovery GWAS used in this study can be found in **Table S2**.

### Genetic architecture of 14 endpoints in GBMI

We first estimated the genetic architecture of 14 endpoints based on HapMap3 SNPs (see STAR Methods). Different prediction methods vary in which SNPs are selected and which effect sizes are assigned to them. Thus, understanding the genetic architecture of complex traits along with sample size and ancestry composition of the discovery GWAS is critical for choosing optimal prediction methods. For example, the SNP-based heritability 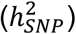 bounds PRS accuracy. We used SBayesS^20^ to estimate 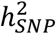, polygenicity (the proportion of SNPs with nonzero effects), and the relationship between minor allele frequency (MAF) and SNP effects (i.e., a metric of negative selection, hereafter denoted as S) for the 14 endpoints in GBMI. Meta-analyses in GBMI were performed across up to 18 different biobanks on 14 endpoints using an inverse-variance weighted method as described in Zhou et al.^18^, including individuals from diverse ancestries. In addition to presenting results using EUR only GWAS summary statistics (EUR GWAS), we also reported estimates using meta-analysis from all ancestries (multi-ancestry GWAS). We explored whether we can reasonably use EUR-based LD reference to approximate the LD of multi-ancestry GWAS in GBMI using the attenuation ratio statistic estimated from LD score regression (LDSC) (see STAR Methods). The attenuation ratio can be used to quantify whether there was a strong LD mismatch, for which the values > 0.2, between GWAS summary statistics and the LD reference panel^21^. We found that the ratio of LDSC using the EUR LD reference panel for GBMI multi-ancestry GWAS was not statistically larger than 0.2. Also, the values were not statistically different from those achieved using GBMI EUR GWAS. This is consistent with a previous study which has found that EUR-based LD can reasonably approximate the LD in their multi-ancestry GWAS consisting of ∼75% EUR individuals^22^.

Most diseases analyzed here had low but significant 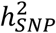 and a range of polygenicity estimates (**Figure 2**). Note that here we reported the 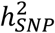 on the liability scale (see STAR Methods). The SBayesS model failed to converge for HCM, likely because its estimated 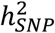 was found to be not significantly different from 0 using LDSC. This could be ascribed to its known predisposing monogenic mutations, the low disease prevalence and heterogeneous subtypes^19^. Therefore, this endpoint was dropped from downstream analyses. We observed that the estimates were overall higher using multi-ancestry GWAS compared to EUR GWAS (**Figure 2**). Overall, the median estimates of SNPs with nonzero effects across 13 endpoints were 0.34% for multi-ancestry GWAS and 0.14% for EUR GWAS (*p*-value = 0.002, paired wilcoxon signed rank test), respectively. The corresponding median estimates for 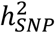 were 0.051 for multi-ancestry GWAS and 0.043 for EUR GWAS (*p*-value = 0.002, paired wilcoxon signed rank test), respectively. The largest difference of 0.06 was found in gout. This could be due to higher 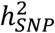 estimated in non-EUR GWAS. For example, the estimates for 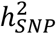 using EUR and EAS GWAS was 0.051 (s.e. = 0.0027) and 0.088 (s.e. = 0.005), respectively. Moreover, we have also found that the estimated effect sizes of two gout-associated loci (close to genes *ALDH16A1* and *SLC2A9*) were different across ancestries^19^. Specifically, we observed that a few top gout-associated variants showed much higher allele frequencies in EAS as compared to EUR, thus resulting in larger variance explained (**Figure S1**).

**Figure 2.**
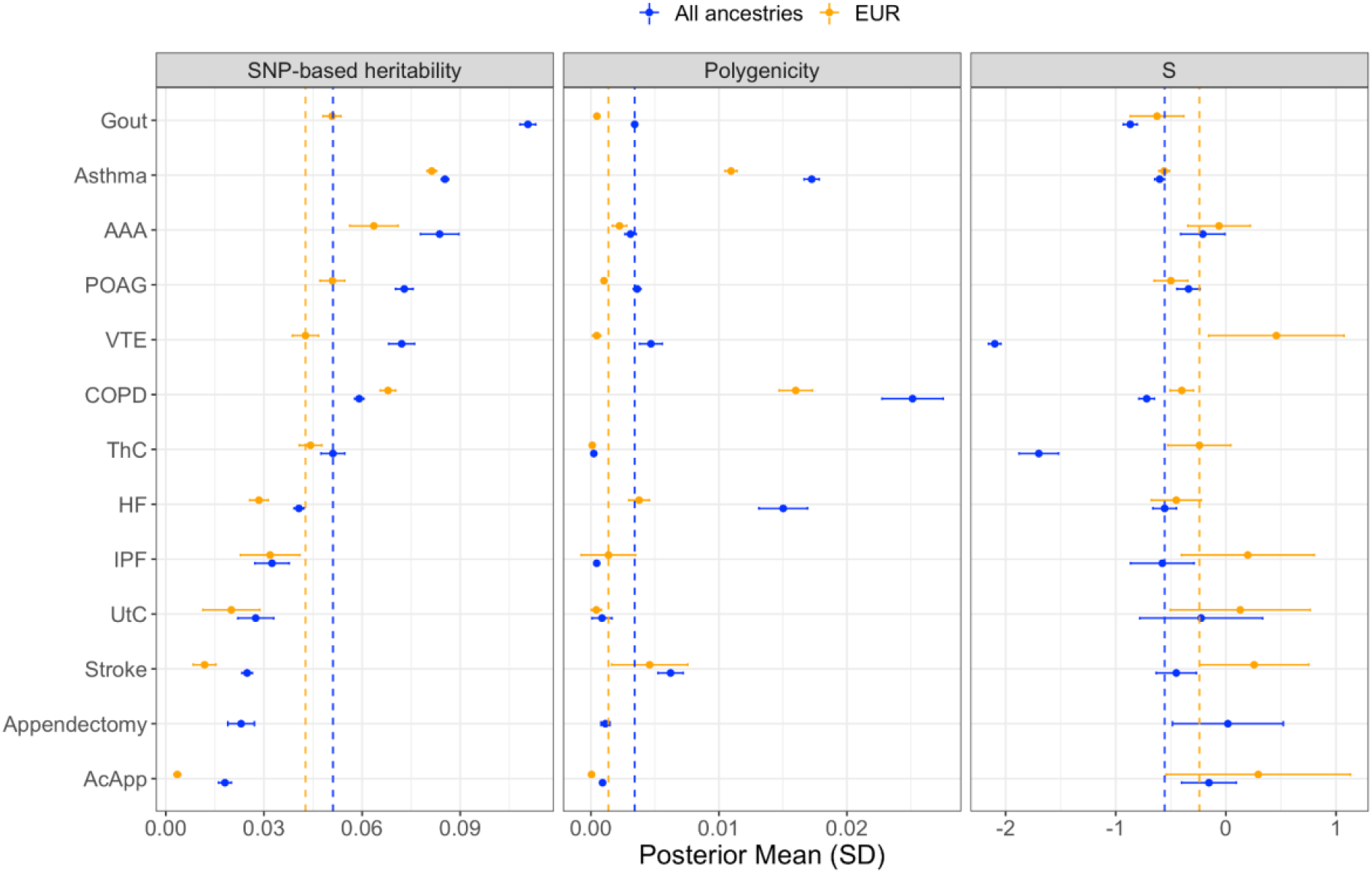
Genetic architecture of endpoints in GBMI. We reported the estimates from using meta-analyzed GWAS from all ancestries (labeled as All ancestries) and European only (labeled as EUR), respectively. The phenotypes on the y-axis are ranked based on the SNP-based heritability estimates using meta-analysis from all ancestries. Note the SNP-based heritability estimates were transformed on the liability scale. The vertical dashed lines in each panel indicate the corresponding median estimates across 13 endpoints. The results for hypertrophic or obstructive cardiomyopathy (HCM) are not presented. Abbreviations: Europeans (EUR), chronic obstructive pulmonary disease (COPD), heart failure (HF), acute appendicitis (AcApp), venous thromboembolism (VTE), primary open-angle glaucoma (POAG), uterine cancer (UtC), abdominal aortic aneurysm (AAA), idiopathic pulmonary fibrosis (IPF), thyroid cancer (ThC).

Polygenicity and 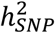 estimates varied greatly among different endpoints. Specifically, the 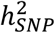 estimates were highest for asthma and gout using multi-ancestry GWAS (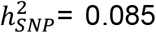 and 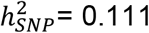, s.e. = 0.0024 respectively), while asthma was found to be much more polygenic than gout. We caution that the numeric interpretation of polygenicity depends on various factors and cannot be interpreted as the number of causal variants. For example, larger and more powerful GWAS tend to discover more trait-associated variants, thus appear to have higher polygenicity. Because we used the same set of SNPs in SBayesS analyses for all endpoints, we hence used the results as a relative measurement of the degree of polygenicity. We observed that the estimate of polygenicity for UtC using multi-ancestry GWAS was not statistically different from 0 (Wald test, *p*-value > 0.05/13) due to limited power observed as relatively low 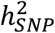. Overall, COPD and asthma were estimated to be the most polygenic traits, followed by HF and stroke, whereas AcApp, UtC and ThC were the least polygenic. Lastly, we observed signals of negative selection for traits including asthma (S = -0.56, s.e. = 0.05), COPD (S = -0.40, s.e. = 0.11) and POAG (S = -0.50, s.e. = 0.15) when considering using EUR GWAS, consistent with empirical findings of negative selection explaining extreme polygenicity of complex traits^23^.

In summary, we observed largely varied key parameters of genetic architecture among 13 endpoints using multi-ancestry and EUR only GWAS. We found that asthma and COPD had the highest 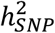 as well as polygenicity. We excluded HCM in our subsequent prediction analyses due to lower evidence of polygenicity and its non-significant 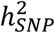.

### Optimal prediction performance using heuristic methods depends on phenotype-specific genetic architecture

We first evaluated the pruning and thresholding (P+T, *p*-value thresholds ranged from 5 × 10^−8^ to 1) method using the EUR-based LD reference panel for all 13 endpoints in the UKBB and BBJ, respectively, given its widespread use and relative simplicity. Note in this study, we used leave-one-biobank-out meta-analyzed GWAS as the discovery GWAS when evaluating PRS in that specific biobank (**Table S2**). We further explored how different factors impact the prediction performance of P+T in diverse ancestry groups, including LD parameters (LD window sizes and LD *r*^2^ thresholds), LD reference panels (ancestry composition, sample size, and SNP density) and per-variant effective sample size (*N*_*eff*_) and MAF (see STAR Methods).

First of all, we selected the optimal *p*-value threshold (the *p*-value threshold with highest prediction accuracy, as measured by *R*^2^ on the liability scale, 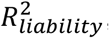, if not specified) in the tuning cohorts and evaluated the accuracies in the test cohorts (see STAR Methods). Specifically, we found that for UKBB with diverse ancestries, using ancestry-specific tuning cohorts provided better prediction performance as compared to that using EUR-based tuning cohorts (**Figure S2**). We found that the optimal *p*-value threshold differed considerably between various endpoints (**Figure S3 and Table S3**). This pattern is found to be related to polygenicity of studied endpoints; but it is also due to a combination of factors such as the GWAS discovery cohort sample size, disease prevalence, trait-specific genetic architecture, and genetic and environmental differences between discovery and target ancestries^24^. For example, when the optimal *p*-value was determined in the UKBB-EUR subset, the less polygenic traits of ThC (106 variants) and AcApp (17 variants) showed highest accuracy at *p*-value thresholds of 5 × 10^−5^ and 5 × 10^−7^, respectively, while for the more polygenic traits of stroke (115,609 variants), HF (115,741 variants), asthma (7,858 variants) and COPD (29,751 variants) achieved the highest accuracy when including SNPs with *p*-value less than 1, 1, 0.01 and 0.1, respectively. To investigate whether ancestries affect the optimal *p*-value threshold, we replicated our analysis in the BBJ (**Figure S3**). In the BBJ, *p*- value thresholds of 5 × 10^−5^, 0.01 and 5 × 10^−5^ presented best performance for gout, stroke and HF, respectively. Consistent with previous studies, these results suggest that optimal prediction parameters (here *p*-value threshold specifically) for P+T appear to be dependent on the ancestry of the target data among other factors25,26. Further, we found that for more polygenic traits including asthma, COPD, stroke and HF, prediction was more accurate with more variants in the PRS (i.e., a less significant threshold) than using the genome-wide significance threshold (*p*-value < 5 × 10-8). On the contrary, less polygenic traits showed no or modest improvement with less stringent *p*-value thresholds, especially for traits such as gout which has trait-associated SNPs with large effects. However, these trends were less obvious in the BBJ which might be attributed to the small proportion of EAS included in the discovery GWAS. One caveat we noted was that fixed LD parameters of P+T were used, thus the results might be impacted by additional optimization of those parameters, which we will further explore below.

We found that further optimizing LD parameters, including LD window size and LD *r*^2^ thresholds, of P+T did not contribute to significant improvement of accuracy across endpoints. Specifically, we observed that the median accuracies with versus without LD parameter optimization were of 0.018 and 0.015, respectively (**Figure S4**). However, there was slight but statistically significant accuracy improvement in EUR for asthma (∼0.006). This might be due to more stratified signals being tagged, which results in noise reduction of the predictor. As compared to using fixed LD parameters, we found similar relationships between polygenicity and optimal *p*-value thresholds when optimizing LD parameters in the UKBB. Specifically, the optimal *p*-value thresholds were overall less stringent for more polygenic traits and more stringent for less polygenic traits. For example, the accuracy using LD parameter optimization in the UKBB-EUR was highest with the *p*-value thresholds of 0.5, 1, 0.1 and 0.2 for the highly polygenic traits of stroke, HF, asthma and COPD, respectively. In contrast, the optimal *p*-value thresholds of 5 × 10^−5^ and 5 × 10^−7^ were observed for less polygenic traits of ThC and AcApp, respectively. To balance the computational burden and signal-to-noise ratio, we used an LD window size of 250Kb and LD *r*^2^ of 0.1 as before. We repeated our analyses using genome-wide common SNPs and compared the prediction accuracy with that using HapMap3 SNPs only (**Figure S4 and Table S3**). There were no significant improvements in prediction accuracies using a denser SNP set, which suggests that HapMap3 SNP set represents genome-wide common SNPs well. Specifically, we found the accuracies in EUR for the most polygenic traits, asthma (∼0.006), COPD (∼0.005) and HF (∼0.004), to be slightly improved using HapMap3 SNPs. Moreover, we found that the sample sizes of the LD reference panel had little impact on P+T performance (**Figure S5**); but the parameters described above including LD window sizes and LD *r*^2^ thresholds had a larger impact on accuracy. We also showed that using 1KG-EUR as the LD reference panel performed well compared to using other ancestral populations with similar sample sizes in the 1KG dataset, which could be explained by the overrepresentation of EUR participants (∼76.4%) in GBMI (**Figure S6 and Table S3**). We further ran LDSC using the EUR-based LD reference panel on leave-specific-biobank-out GWAS in GBMI to estimate the attenuation ratio statistic (see STAR Methods). Similar to previous findings, we found that even in leave-UKBB-out GWAS with the lowest EUR proportion (**Table S2**), its LD information can be well approximated using the EUR reference panel, which was reflected by the values of ratio not statistically larger than 0.2 and not statistically different from EUR GWAS in GBMI. We therefore used 1KG-EUR as the LD reference panel for all subsequent P+T analyses. But the choice of external LD reference panel for multi-ancestry GWAS needs further exploration especially when the discovery GWAS becomes more diverse.

Finally, we investigated the impact of per-variant effective sample size heterogeneity. Since GBMI consists of a number of biobanks with diverse ancestries, the number of samples used for meta-analysis was notably heterogeneous among the variants; the majority of the variants in the GWAS meta-analysis had only a limited number of effective samples (*N*_*eff*_) (**Figure 3-A**). Therefore, although sample size heterogeneity is not usually considered for PRS, it may confound the PRS prediction accuracy in the case of global biobank collaborations. By filtering the variants according to *N*_*eff*_ per- variant (i.e., *N*_*eff*_ larger than 50% or 80% thresholds of the maximum *N*_*eff*_ of the trait of interest, see STAR Methods), we observed that the 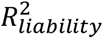 increased substantially for less stringent thresholds (*p*-value > 5 × 10^−5^) in the UKBB (**Figure S7-A**). As a representative example, the largest 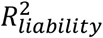 (0.034) was obtained for asthma when the *p*-value threshold was 5 × 10^−3^, whereas the 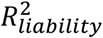 was 6.6 × 10^−3^ at the threshold without *N*_*eff*_ filtering (**Figure 3-B and Table S4**). Next, we investigated whether *N*_*eff*_ filtering could be substituted by other filtering criteria. Although excluding variants with MAF less than 0.1 partially compensated for PRS transferability, the improvement of *N*_*eff*_ filtering in 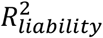 was still observed (**Figure S7-B**). Heterogeneity in *N*_*eff*_ might be confounding especially in multi-ancestry meta-analyses because it can be distorted by heterogeneous allele frequencies and imputation quality spectra among ancestries. Indeed, as rarer variants tend to be more ancestry-specific, variants with low *N*_*eff*_ tend to be unique to specific ancestries (**Figure 3-C**). Of note, the dependency of 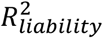 on the *N*_*eff*_ was, however, largely rectified for most of the traits by using only HapMap3 SNPs (**Figure S7-C**). Given that the 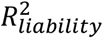 for HapMap3 SNPs was comparable to that for genome-wide SNPs (**Figure S4**), filtering to HapMap3 SNPs might be suitable for meta-analysis of diverse populations. On the other hand, HapMap3 SNPs generally have good imputation quality, although a recent study shows that relaxing imputation INFO score from 0.9 to 0.3 has negligible impacts on prediction accuracy^9^. We replicated the *N*_*eff*_ filtering in BBJ and confirmed that improved 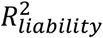 attributable to *N*_*eff*_ filtering was also observed (**Figure S7-D**). Although the effect of the *N*_*eff*_ filtering was diminished by the MAF filtering in relatively stringent thresholds (*p*-value < 5 × 10^−4^), the effect was still observed in the other thresholds (**Figure S7-E**). Using only HapMap3 SNPs almost completely reduced the dependency of 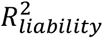 on the *N*_*eff*_ (**Figure S7-F**).

**Figure 3.**
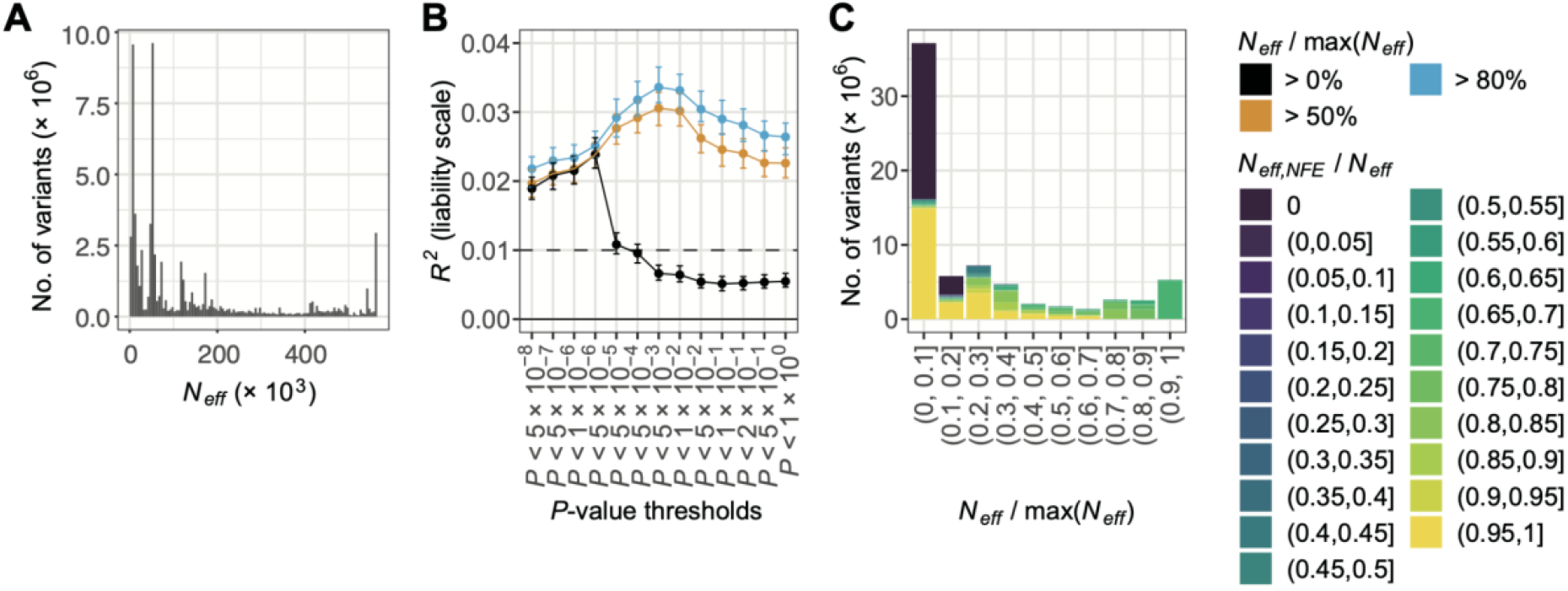
Sample size heterogeneity affects PRS prediction accuracy for P+T. A) the distribution of effective sample sizes (*N*_*eff*_) for asthma as a representative trait. B) predictive performance of P+T for European (EUR) samples in the UK Biobank (UKBB). The *R*^*2*^ for asthma is shown as a representative result. Full results are shown in Figure S7 and Table S3. C) the ratio of *N*_*eff*_ of EUR compared with *N*_*eff*_ of all samples for asthma.

Overall, we found the prediction performance of P+T to be affected by a combination of factors, with *p*-value thresholds showing larger effects as compared to other parameters, such as LD window sizes, LD *r*^2^ thresholds, and variant filtering by *N*_eff_ or MAF. Moreover, the optimal *p*-value threshold varied substantially between different endpoints in GBMI. We also demonstrated that restricted use of HapMap3 SNPs showed comparable or better prediction accuracy relative to using genome-wide common SNPs for P+T, particularly for GWAS from diverse cohorts as in GBMI with genetic variants showing considerable heterogeneity in effective sample sizes.

### Bayesian approaches for calculating PRS improve accuracy

We also evaluated fully genome-wide polygenic risk scores, by first fine-tuning the parameters in PRS-CS. We ran PRS-CS using both the grid model and automated optimization model (referred to as auto model), the former of which specifies a global shrinkage parameter (phi, in which smaller values indicate less polygenic architecture and vice versa for larger values), with 1KG- EUR as the LD reference panel. We note that the optimized phi parameter with highest prediction accuracy in the grid model differed among traits (**Figure S8**). Specifically, we found that for more polygenic traits (as estimated using SBayesS) including asthma, COPD and stroke (**Figure 2**), the optimal phi parameter was 1 × 10^−3^ in EUR (**Figure S8**). There was no significant difference between prediction accuracy using the optimal grid model versus auto model (**Figure S8**), which suggests PRS-CS can learn the phi parameter from discovery GWAS well when its sample size is considerably large. Therefore, we hereafter used the auto model because of its computational efficiency. Across target ancestral populations in the UKBB, PRS from EUR-based LD reference panels showed significantly higher or comparable prediction accuracies compared to PRS using other ancestry-based LD reference panels (**Figure S9-A**). This result suggests that it is reasonable to use a EUR-based LD reference panel in GBMI largely because EUR ancestry constitutes the largest proportion of GWAS participants (∼76.4%). Note that we also compared the prediction accuracy of LD reference panels derived from UKBB-EUR, which has a much larger sample size, against 1KG-EUR and found no significant difference (**Figure S9-B**). These results suggest that PRS-CS is not sensitive to the sample size of the LD reference panel, which is consistent with previous findings^27^.

We then compared the optimal prediction accuracy of P+T versus the PRS-CS auto model in the UKBB and BBJ and found that PRS-CS showed overall better prediction performance for traits with higher 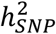 but no or slight improvements for traits with lower 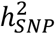 (**Figure 4**). Specifically, the highest significant improvement of PRS-CS relative to that of P+T in EUR was observed for HF, of 60.9%, followed by COPD (53.2%) and asthma (48.8%). Substantial increments were observed for HF (105.2%), COPD (102.5%) and asthma (60.9%) in EAS. 45.8% and 48.1% improvements were shown for asthma in CSA and AFR, respectively. P+T saw better prediction performance over PRS-CS for a few trait-ancestry comparisons, however, such improvement was not statistically significant. Compared with P+T, which requires tuning *p*-value thresholds and is affected by variant-level quality controls such as *N*_*eff*_, there is no need to tune prediction parameters using the PRS-CS auto model, thus reducing the computational burden.

**Figure 4.**
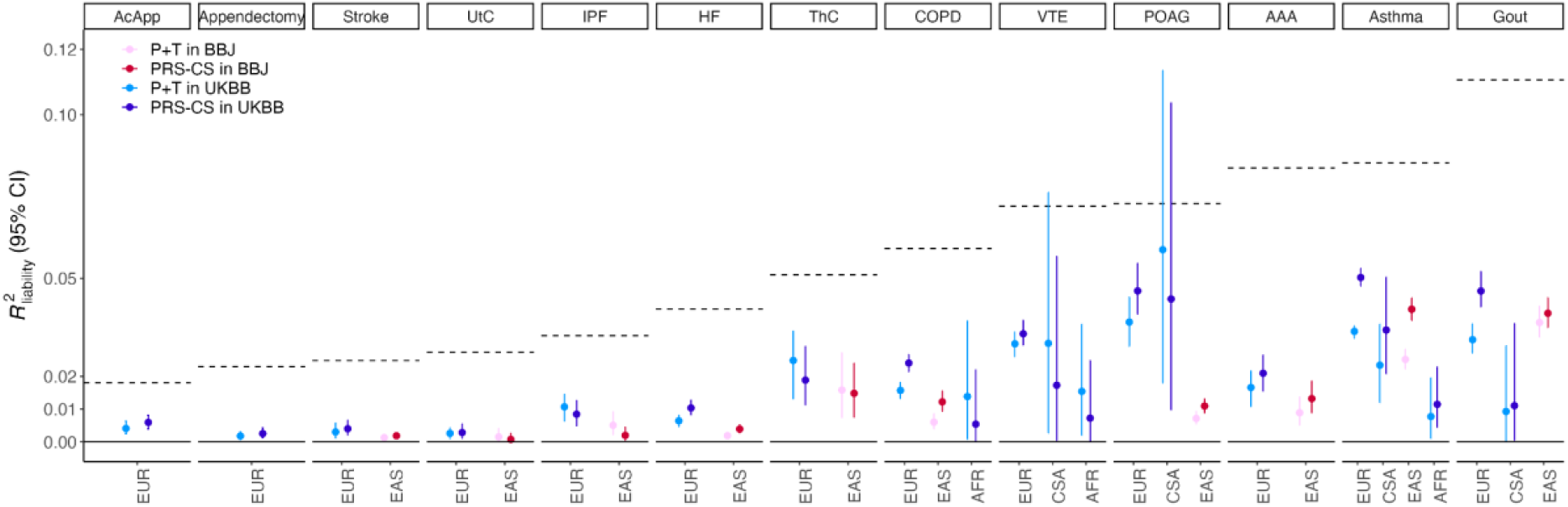
Prediction performance using P+T versus that using PRS-CS. The phenotypes are ranked based on the SNP-based heritability as shown in **Figure 2** (indicated by the dashed line) estimates using all ancestries. Only trait-ancestry pairs with significant accuracies in both P+T and PRS-CS are presented. The prediction accuracy in P+T was estimated in the test cohort based on the optimal *p*-value thresholds fine- tuned in the validation cohort. The auto model was used for PRS-CS. Abbreviations: Europeans (EUR), Admixed Americans (AMR), Middle Eastern (MID), Central and South Asians (CSA), East Asians (EAS) and Africans (AFR), chronic obstructive pulmonary disease (COPD), heart failure (HF), acute appendicitis (AcApp), venous thromboembolism (VTE), primary open-angle glaucoma (POAG), uterine cancer (UtC), abdominal aortic aneurysm (AAA), idiopathic pulmonary fibrosis (IPF), thyroid cancer (ThC).

Overall, after examining 13 disease endpoints, these results favor the use of PRS-CS for developing PRS from multi-ancestry GWAS of primarily European samples, which is also consistent with previous findings that Bayesian methods generally show better prediction accuracy over P+T across a range of different traits^9,27^. The practical considerations about the two models, PRS-CS and P+T, used in this study, are shown in **Table S5**.

### PRS accuracy is heterogeneous across ancestries and biobanks

For each of the participating biobanks, we used leave-one-out meta-analysis as the discovery GWAS to estimate the prediction performance of PRS in each biobank (see STAR Methods). The disease prevalence and effective sample size of each biobank is shown in **Figure S10**. Generally, the PRS prediction accuracy of different traits increased with larger 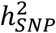 (**Figure 5 and Table S6**). For example, the average *R*^2^ on the liability scale across biobanks (hereafter denoted as 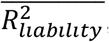, see STAR Methods) in EUR ranged from <1.0% for AcApp, appendectomy, stroke, UtC and IPF, 1.0% for HF, ∼2.2% for COPD and ThC to 3.8% for gout and 4.6% for asthma. Notably, accuracy was sometimes heterogeneous across biobanks within the same ancestry for some traits. Specifically, the 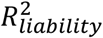 for asthma in ESTBB and BioVU was significantly lower than 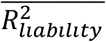, which might be attributable to between-biobank differences such as recruitment strategy, phenotyping, disease prevalence, and environmental factors. The prediction accuracy was generally lower in non-European ancestries compared to European ancestries, especially in African ancestry, which is mostly consistent with previous findings^28–30^ with a few exceptions. For example, we observed comparable prediction accuracy for gout in EAS relative to that in EUR, which could be reflected by large effective sample sizes and some gout-associated SNPs with large effects exhibiting higher allele frequencies in EAS (**Figure S1**). For example, the MAFs of gout top-associated SNP, rs4148157, were 0.073 in 1KG-EUR and 0.25 in 1KG-EAS, respectively, and the phenotypic variance explained by that SNP in EAS (8.3%) was more than twice as high as that in EUR (3.0%). The accuracy of PRS to predict asthma risks in AMR was found to be significantly higher than that in EUR, which could be due to the small sample size in AMR (**Table S6**). Thus, further validation is needed in larger AMR population cohorts.

**Figure 5.**
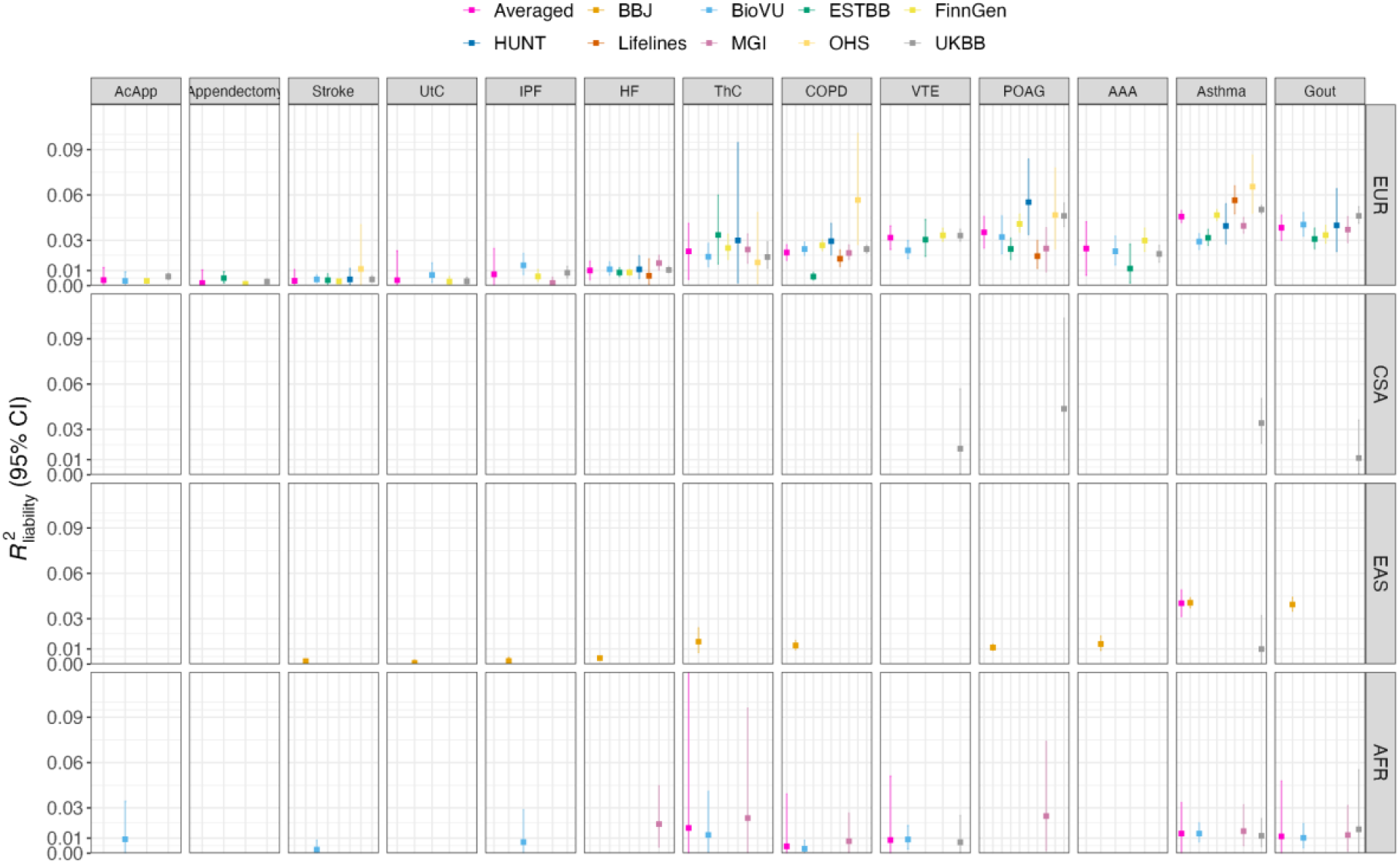
Prediction performance of PRS-CS across biobanks and ancestries. The phenotypes on the y-axis were ranked by the SNP-based heritability using all ancestries as shown in Figure 2. Only the significant results were shown. Data for all trait- ancestry pairs in each biobank are provided in **Table S6**. Note that we removed the estimates in AMR and MID due to limited information as a result of small sample sizes. Abbreviations: Europeans (EUR), Admixed Americans (AMR), Middle Eastern (MID), Central and South Asians (CSA), East Asians (EAS) and Africans (AFR), chronic obstructive pulmonary disease (COPD), heart failure (HF), acute appendicitis (AcApp), venous thromboembolism (VTE), primary open-angle glaucoma (POAG), uterine cancer (UtC), abdominal aortic aneurysm (AAA), idiopathic pulmonary fibrosis (IPF), thyroid cancer (ThC).

The ability of PRS to stratify individuals with higher disease risks was also found to be heterogeneous across biobanks and ancestries as shown in **Figure 6** and **Table S7**. We showed that the PRS distribution across different biobanks slightly varied. Specifically, we calculate d the absolute difference of median PRS in each decile for each endpoint between biobanks for cases and controls, separately, and found that the largest absolute differences were 0.06 and 0.21 for stroke controls and stroke cases, respectively (**Figure S11**). This justifies the comparison of odds ratios (ORs) in terms of relative risks. The ORs between the top 10% and bottom 10% were more heterogeneous between biobanks and also higher relative to other comparisons (e.g., top 10% vs middle and other strata). This is consistent with previous studies where OR reported between tails of the PRS distribution is generally inflated relative to those between top ranked PRS and general populations^11^. We measured the variation of OR between biobanks using the coefficient of variation of OR (CoeffVar_OR_, see STAR Methods). The largest CoeffVar_OR_ in EUR was observed for ThC of 0.46 between top 10% and bottom 10% as compared to 0.27 and 0.23 for top 10% vs middle and other, respectively. We recapitulated the findings using 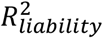 that ORs were overall higher for traits with higher 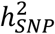 and also higher in EUR than non-EUR ancestries, which is expected as the two accuracy metrics are interrelated. For example, the averaged ORs across biobanks weighted by the inverse variance in EUR (see STAR Methods) for gout were 4.6, 2.4 and 2.2 for the top 10% vs bottom 10%, middle and other strata, separately. The corresponding estimates in EUR for stroke were 1.6, 1.3 and 1.3, respectively. Across ancestries, the average OR of asthma between the top 10% and bottom 10% ranged from 4.1 in EUR to 2.4 in AFR.

**Figure 6.**
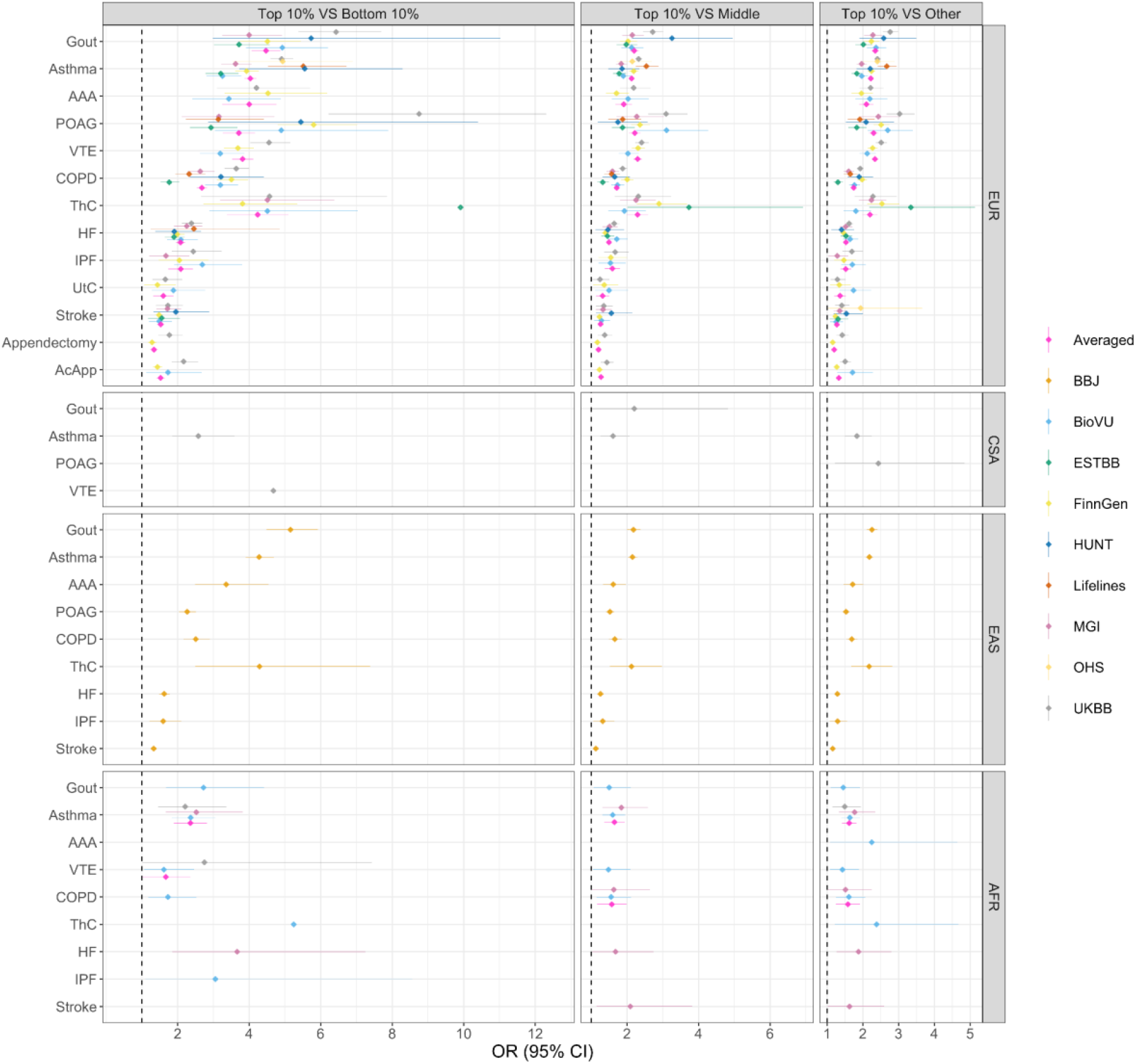
The odds ratio (OR) between different PRS strata for endpoints in GBMI. The dashed line indicates OR=1. Only significant trait-ancestry specific OR was reported, with p-value < 0.05. The full results are shown in Table S7. The averaged OR was calculated using the inverse-variance weighted method (see STAR Methods). PRS was stratified into deciles with the first decile (bottom 10%) used as the referenced group. The phenotypes were ranked based on SNP-based heritability estimates using all ancestries (see Figure 2). Abbreviations: Europeans (EUR), Admixed Americans (AMR), Middle Eastern (MID), Central and South Asians (CSA), East Asians (EAS) and Africans (AFR), chronic obstructive pulmonary disease (COPD), heart failure (HF), acute appendicitis (AcApp), venous thromboembolism (VTE), primary open-angle glaucoma (POAG), uterine cancer (UtC), abdominal aortic aneurysm (AAA), idiopathic pulmonary fibrosis (IPF), thyroid cancer (ThC).

Overall, the predictive performance of PRS measured by 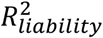 and OR was found to be heterogeneous across ancestries. This heterogeneity was also presented across biobanks for traits such as asthma which is considered as a syndrome comprising heterogeneous diseases^31^.

### GBMI facilitates improved PRS accuracy compared to previous studies

GBMI resources might be expected to improve prediction accuracy due to large sample sizes and the inclusion of diverse ancestries. To explore this, we compared the prediction accuracy achieved by GBMI versus previously published GWAS using the same pipeline to run PRS-CS. As shown in **Figure 7 and Figure S12**, the accuracy improvements were most obvious for traits with larger 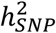 but there was no or slight improvement for traits with lower 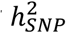. Specifically, we calculated the absolute improvement of GBMI relative to that using previously published GWAS and found that on average across biobanks, the largest improvements of 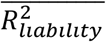 in EUR were 0.033 for asthma, 0.031 for gout, 0.019 for ThC and 0.017 for COPD, whilst the corresponding improvements of AUC on average 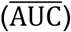 were 0.051, 0.078, 0.078 and 0.041, respectively. Substantial improvements were also observed for gout in EAS 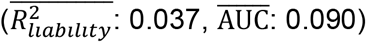, for asthma in CSA 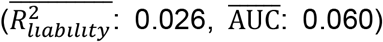, 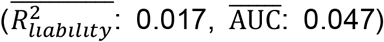 and AFR 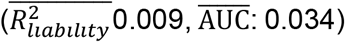, and for ThC in EAS 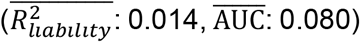 and AFR 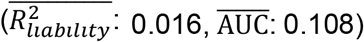. However, PRS accuracy was significantly higher for published GWAS relative to the current GBMI for POAG in EUR and AFR, and COPD in the specific case of Lifelines biobank. We referred to the datasets included in the public GWAS of POAG and found that individuals from diverse datasets of EUR and AFR populations were also part of the discovery dataset, thus we cannot rule out the possibility of sample overlapping or relatedness between the discovery and target datasets for these populations. This suggests that the PRS evaluation may be biased upwards from the prior GWAS for POAG. Also, the phenotypes of POAG across different biobanks are likely more heterogeneous in GBMI than targeted case-control studies^18,32^. The meta-analysis of GBMI with International Glaucoma Genetics Consortium (IGGC) did not lead to substantially improved prediction performance^32^. Another concern might be the disproportional case/control ratio of POAG in GBMI, of ∼27,000 cases and ∼1.4M controls, thus POAG-related phenotypes with shared genetics in the controls or possible uncontrolled ancestry differences between cases and controls might confound the GBMI GWAS. A very high heterogeneity for phenotype definitions is also found for COPD, however this does not explain why one biobank alone presents this pattern; a specific environmental or population effect not considered in the broad analysis might affect this particular observation.

**Figure 7.**
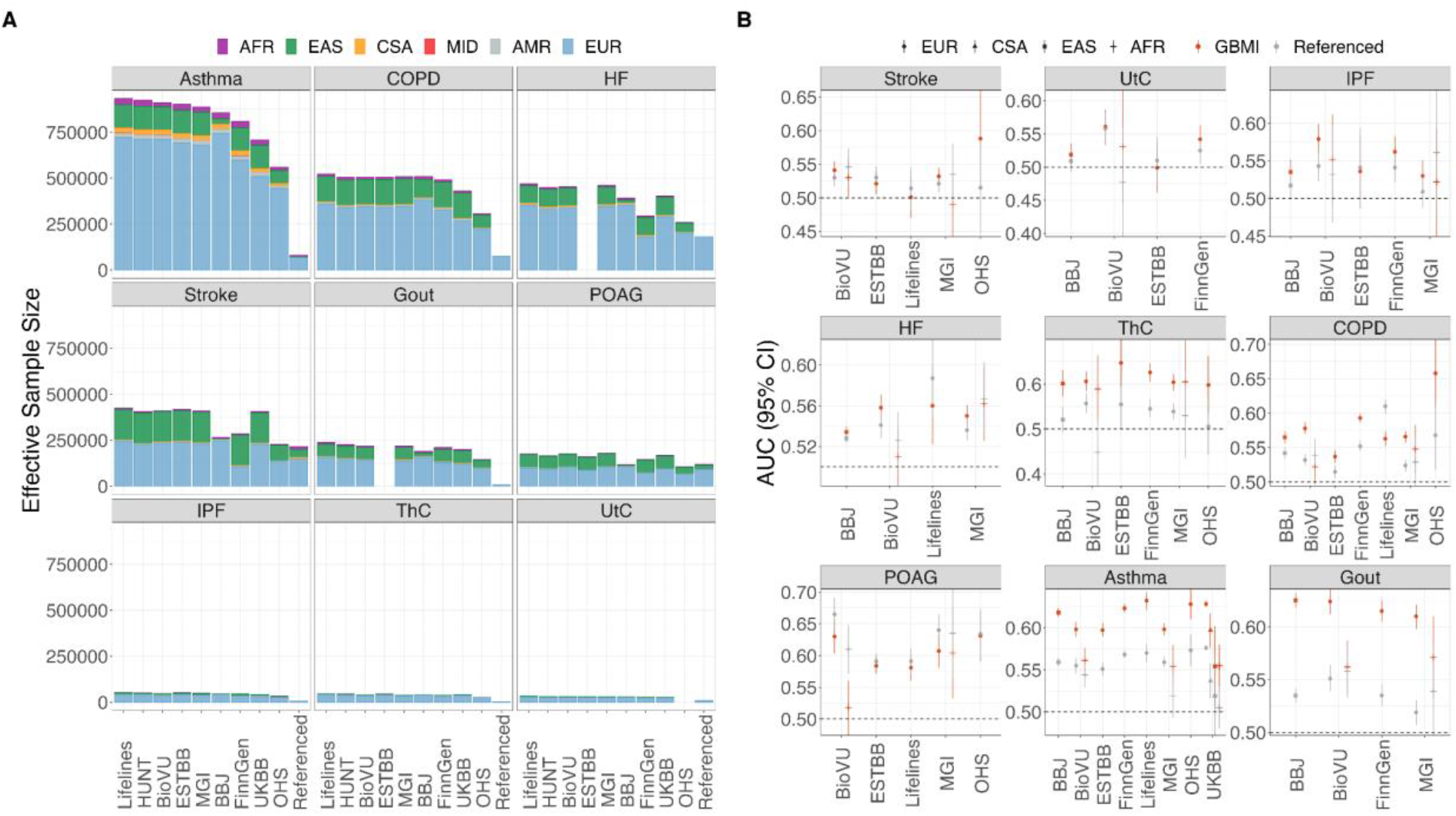
The prediction performance and ancestry compositions of GBMI versus previously published GWAS. A) The ancestry compositions of GBMI and referenced GWAS. The label for biobanks in the x-axis indicated the leave-one-out-biobank meta-analyzed GWAS in GBMI. The previously published GWAS was labeled as Referenced. B) The comparison of AUC between GBMI and referenced GWAS. The AUC was calculated by fitting PRS only. The phenotypes in A) were ranked based on the effective sample sizes from all ancestries. The phenotypes in B) were ranked by the SNP-based heritability estimates from all ancestries. Note that we removed the estimates in AMR and MID due to limited information as a result of small sample sizes. The full results are shown in **Table S4**. Abbreviations: Europeans (EUR), Admixed Americans (AMR), Middle Eastern (MID), Central and South Asians (CSA), East Asians (EAS) and Africans (AFR), chronic obstructive pulmonary disease (COPD), heart failure (HF), acute appendicitis (AcApp), venous thromboembolism (VTE), primary open-angle glaucoma (POAG), uterine cancer (UtC), abdominal aortic aneurysm (AAA), idiopathic pulmonary fibrosis (IPF), thyroid cancer (ThC).

To boost statistical power, we can meta-analyze GBMI GWAS with other non-overlapping cohorts as shown in other GBMI working groups^33–35^. However, we should note that more heterogeneity might be introduced from different resources such as population structure and phenotype definitions, which we cannot control with summary statistics data and that could exacerbate the heterogeneous performance of PRS across target populations. On the other hand, GBMI is open to more cohorts and has been continuously working on integrating more datasets.

## Discussion

The GBMI resource is notable in its collection of phenotypes studied and range of participating cohorts from multiple ancestry groups; it has therefore offered a unique opportunity to comprehensively evaluate and develop guidelines regarding the effects of multi-ancestry and heterogeneous GWAS discovery data, polygenicity, and PRS methods on prediction performance in diverse target cohorts. In this study, we have used the unique GBMI resource consisting of multi-ancestry GWAS for multiple disease endpoints with varying genetic architectures and prevalences across diverse populations to develop and evaluate PRS. Indeed, we found overall across a range of phenotypes and ancestries that using the large-scale meta-analysis from GBMI significantly improved PRS accuracy compared to previous studies with smaller sample sizes and less diverse cohorts. While some previous studies have benchmarked PRS methods and accuracies, most have been based on relatively homogeneous GWAS discovery cohorts or evaluated for specific phenotypes^3,9,26,36^. Even when assessing the portability of PRS across ancestries, most evaluations have included ancestrally diverse target cohorts but still relatively homogeneous discovery cohorts^12,13,37^. Thus, based on the results of our analyses using GBMI, we have provided additional lessons and guidelines for developing PRS with multi-ancestry discovery data for different endpoints (**Figure S13**). We have organized these best practices according to 1) characteristics of the discovery GWAS, 2) PRS model fitting, and 3) the target cohort.

First, the GWAS discovery cohort provides the prerequisite input for polygenic score calculations and interpretation, namely how phenotypes are ascertained and in which populations, which SNPs to include, and which effect sizes will be used. We recommend that standard quality controls should be performed with more caution when considering multi-ancestry discovery GWAS. Specifically, we suggest filtering variants based on the per-variant effective sample size (*N*_*eff*_) and MAF as they show considerable heterogeneity across datasets and ancestries in our discovery GWAS. When we filtered out variants with extremely small *N*_*eff*_ in our P+T analyses, and in particular when using HapMap3 SNPs, PRS prediction performance improved. As noted in Zhou et al.^19^, the allele frequencies of variants in GBMI meta-analyzed GWAS were compared with those in gnomAD using Mahalanobis distance and flagged if they were three standard deviations away from the mean. We recommend computing such statistics and filtering with this information, or if infeasible, restricting to using only HapMap3 variants.

Given the significant improvements in PRS accuracy with GBMI discovery GWAS over previous studies with smaller sample sizes and less diversity, we recommend using the largest and most diverse GWAS discovery cohort available when constructing PRS, even if it matches the ancestry composition of the target cohort slightly less well than a smaller GWAS. Overall, traits with higher SNP-based heritability showed greater improvement compared to those with lower SNP-based heritability. This indicates that PRS performance will continually benefit from larger sample sizes and more diverse populations. However, further research is needed to understand more concretely how the composition of underrepresented populations, including specific ancestries and varying sample sizes, can be modeled alongside current Eurocentric GWAS to best facilitate PRS accuracy and generalizability.

Second, when fitting PRS models, important choices include which PRS construction methods to use, how to fine-tune hyperparameters, and which LD reference panels to use. So far, PRS models that use GWAS summary statistics have been favored over those that use individual-level data due to their computational efficiency and data access restrictions. These models have been comprehensively reviewed recently^10,38^. In this study, we therefore explored the prediction performance of two widely used PRS construction methods, P+T and PRS-CS. We paired the results of these methods with prior knowledge of trait-specific genetic architecture estimates from SBayesS. The best predictor for P+T is often obtained by fine-tuning the *p*-value thresholds in a validation dataset, while other LD related parameters, such as LD *r*^2^ and LD window size, are usually arbitrarily specified. Here, we found that the prediction accuracy of P+T was much less sensitive to different LD-related parameters compared to various *p*-value thresholds. Moreover, the optimal *p*-value threshold varied across phenotypes, likely because of trait-specific genetic architecture, especially the degree of polygenicity measured by SBayesS. However, differences in discovery GWAS and target dataset such as sample sizes, phenotype definition, disease population prevalence and population characteristics could also contribute to this variation. When analyzing PRS-CS results, we validated a previous finding that the auto model, that does not require post-hoc tuning of the proportion of SNPs with non-zero effects (phi), showed similar prediction performance relative to the more computationally intensive grid model, which requires determining the optimal phi parameter in an independent tuning cohort^27^.

We also recommend using prior knowledge and empirical measurements of the genetic architecture of studied phenotypes to choose specific types of PRS models. In this study, we evaluated the effects of trait-specific genetic architecture on PRS performance using estimates from SBayesS. Generally, traits with higher SNP-based heritability, such as asthma and gout, showed greater improvement with the GBMI discovery data compared to those with lower SNP-based heritability, such as acute appendicitis (AcApp). Trait-specific architecture affected both the choice of method and optimal hyper-parameters. For example, extremely polygenic traits are more suitable for an infinitesimal model or Bayesian models that are adaptive to the trait genetic architecture. The specific model hyper-parameters are also affected by trait genetic architecture. For example, the optimal *p*-value threshold of P+T might be more stringent for less polygenic traits but less stringent for highly polygenic traits.

Another decision point in fitting PRS models is regarding which LD reference panel to use when multi-ancestry GWAS discovery and target populations are available. An in-sample LD reference panel that spans the full discovery cohort is optimal but rarely available. Here, we have shown that EUR-based LD reference panels can reasonably approximate the LD of GBMI GWAS. However, choosing LD reference panels that mirrors the ancestry composition of the discovery GWAS when in-sample LD reference panels are not available is ideal. For convenience, if one ancestry is dominant in the multi-ancestry GWAS, we suggest using that ancestry-matched reference panel. The attenuation ratio statistic estimated from LDSC can further be used as a measure to quantify the degree of LD mismatch between discovery GWAS and LD reference panels^22^. When ancestry proportions are relatively evenly distributed, we and others have found that using LD reference panels with ancestry proportions that match the discovery GWAS could provide better prediction performance especially for less polygenic traits with large effect variants (unpublished work), such as lipid traits^39^. We also found that prediction performance can be improved when using ancestry-matched tuning cohorts for PRS construction to fine-tune hyper- parameters and avoid overfitting, such as P+T and the PRS-CS grid models explored in this study. While other studies have also explored options such as pseudo-validation when no additional tuning cohort is available^40,41^

Third, the practical considerations for target populations involved in PRS analyses are quite consistent between using homogenous GWAS and multi-ancestry GWAS. In this study, we used biobanks with various ancestry compositions and recruitment strategies as the target cohorts^19^. For example, BBJ, BioVU and MGI are hospital-based biobanks whereas others are population-based or have mixed enrollment strategies, which can impact phenotype precision or ascertainment bias and therefore heritability. UKBB, MGI and BioVU have diverse ancestries while others primarily consist of one ancestry (either European or East-Asian participants). The performance of PRS in different target populations can also be affected by the ancestry proportions in the discovery GWAS and precision of phenotype definition aside from biobank-specific factors (e.g., environmental factors), which warrants further exploration. We therefore recommend considering those factors and reporting PRS distribution statistics (e.g., median PRS) and accuracy metrics when benchmarking the prediction performance between different PRS predictors. More reporting standards about PRS models have been well-documented in PGS Catalog^36^.

Related to the target cohorts, we also found that the prediction performance showed great heterogeneity across biobanks and ancestries. Because PRS are only intended to capture genetic factors, other considerations such as environmental exposures and demographic history may impact the predictive power of PRS within and across ancestries, with recommendations for how to model these alongside PRS an open question for future research and methods development. For example, we found that the 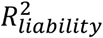 in OHS was overall higher than in other biobanks, which may be attributed to the more complex relatedness structure in this founder population. Notably, the phenotype definitions, recruitment strategy and disease prevalence also vary to different extents across the biobanks studied here.

We note a few limitations in our study. First, we chose 1KG-EUR as the LD reference panel because data security practices often preclude the use of individual-level GWAS data across analytical teams. Although we have shown that the EUR-based LD reference panels can reasonably approximate the LD of GBMI GWAS studied here, it still could affect SNP effect size estimates and thus prediction performance. Further efforts are required to provide more appropriate LD reference panels. For example, utilizing the large-scale UKBB with individual-level genotypes to construct a panel with matched ancestry proportions to the discovery GWAS has been used in a recent study^39^. However, early explorations have shown that using proportional LD reference panels generally achieves similar prediction performance as using EUR-based reference panels when EUR is primarily dominant in the multi-ancestry GWAS (unpublished work). Also, sharing LD matrices from participating biobanks without accessing individual-level data would be another alternative to construct an in-sample LD matrix. On the other hand, individual-level based PRS construction methods across large-scale biobanks without relying on LD reference panels are also promising. Such methods could potentially benefit from secure large-scale GWAS across multiple datasets. For example, Blatt et al.^42^ have used homomorphic encryption to establish a privacy-preserving framework to perform GWAS and decrypt the results for sharing through a project coordinator. Second, we have focused on common SNPs, specifically HapMap3 SNPs for PRS-CS. As a result, information from rarer variants missing in the LD reference panel was not captured in other non-European ancestries, which may explain a small fraction of the loss of accuracy across populations. Third, although a harmonized analysis framework was developed for GBMI, such as phenotype definitions, ancestry assignments, and PRS construction, there remains a multitude of factors that may contribute to heterogeneous accuracy across both biobanks and ancestries. These include, but are not limited to, phenotype precision, cohort-level disease prevalence, and environmental factors. Last, we evaluated PRS predictive performance using multi-ancestry GWAS but comparisons with single-ancestry GWAS at sufficient scale would enable us to better understand the specific contributions of ancestry diversity and increasing sample size especially for under-represented ancestries, which also serves as a future direction.

The GBMI resource constitutes remarkable progress in expanding the number of endpoints and ancestry groups studied, laying the groundwork for several future directions for exploration. For example, PRS construction methods that model GWAS summary statistics alongside LD information from multiple ancestries have shown promising accuracy improvements for some traits^16,43^, but statistical methods are insufficient for equitable accuracy without simultaneous progress in generating large-scale diverse data, as early investigation into one of these methods has yielded marginal improvement in both European and non-European ancestries for asthma in GBMI^44^. In addition to multi-ancestry GWAS, sex-stratified GWAS in GBMI also provides opportunities to explore the role of sex-specific effects as well as impacts from the sample size ratio of males/females on prediction performance of PRS across biobanks. Beyond genetic effects, biobank-specific risk factors and environmental exposures provide further opportunities to better understand the heterogeneity in PRS accuracy that we have identified across biobanks and ancestries^45,46^. This will be extremely important as previous work has shown that prediction performance differences between target cohorts are not likely to be reduced using various PRS construction methods^9^. Finally, extending these collaboration efforts to more biobanks in the future, particularly those including recently admixed populations, will bring more resolution into those effects that are biobank-specific and ancestry-specific. Studies in recently admixed populations show that GWAS power can be improved by utilizing local ancestry-specific SNP effect estimates and thus have the potential to benefit genetic prediction accuracy and generalizability, particularly for less polygenic traits^47,48,49^. Altogether, these initiatives hold great promise for improving transferability of PRS across biobanks and ancestries by harnessing the phenotypic richness and diversity present in different biobanks.

## Supporting information

GBMI_full_authors

## Data Availability

All data produced in the present work are contained in the manuscript

https://www.globalbiobankmeta.org/resources

http://results.globalbiobankmeta.org/

ftp://ftp.1000genomes.ebi.ac.uk/vol1/ftp/data_collections/1000_genomes_project/data

## Acknowledgements

A.R.M is funded by the K99/R00MH117229. E.L. is funded by the Colciencias fellowship ed.783. S.N. was supported by Takeda Science Foundation. Y.O. was supported by JSPS KAKENHI (22H00476), and AMED (JP21gm4010006, JP22km0405211, JP22ek0410075, JP22km0405217, JP22ek0109594), JST Moonshot R&D (JPMJMS2021, JPMJMS2024), Takeda Science Foundation, and Bioinformatics Initiative of Osaka University Graduate School of Medicine, Osaka University. E.R.G. is supported by the National Institutes of Health (NIH) Awards R35HG010718, R01HG011138, R01GM140287, and NIH/NIA AG068026. V.L.F. was supported by the European Union’s Horizon 2020 research and innovation programme under the Marie Skłodowska-Curie grant agreement No.675033 (EGRET plus). L. B. and B. B. receive support from the K.G. Jebsen Center for Genetic Epidemiology funded by Stiftelsen Kristian Gerhard Jebsen; Faculty of Medicine and Health Sciences, NTNU; The Liaison Committee for education, research and innovation in Central Norway; and the Joint Research Committee between St Olavs Hospital and the Faculty of Medicine and Health Sciences, NTNU. K.L. and R.M. were supported by the Estonian Research Council grant PUT (PRG687) and by INTERVENE - This project has received funding from the European Union’s Horizon 2020 research and innovation programme under grant agreement No 101016775. W.Z. was supported by the National Human Genome Research Institute of the National Institutes of Health under award number T32HG010464. The work of the contributing biobanks was supported by numerous grants from governmental and charitable bodies. The biobank specific acknowledgements and full author list for GBMI are included in the **Supplementary Notes**.

## Author Contributions

Study design: A.M., J.H., Y.O., Y.W.

Data collection/contribution: L.B., P.A., B.B., P.D., K.H., R.M., Y.M., S.S., J.U., C.W., N.J.C., I.S., J.H.

Data analysis: Y.W., S.N., E.L., S.K., K.T., K.L., M.K. W.Z., K.H.W, M.J.F., L.B., V.L.F, J.H. Writing: Y.W., S.N., E.L., Y.O., A.M., J.H

Revision: Y.W., S.N, E.L., K.T., W.Z., S.S., J.W.S., B.N.W., C.W., E.R.G., N.J.C., Y.O., A.M., J.H.

## Declaration of Interests

E.R.G. receives an honorarium from the journal Circulation Research of the American Heart Association as a member of the Editorial Board.

## STAR Methods

### Datasets and quality control

Discovery datasets: For each of 14 endpoints, we used GWAS summary statistics from both GBMI and public datasets with summary statistics available in GWAS Catalog if applicable (**Table S1 and Table S2**) as the discovery dataset. We filtered out SNPs with ambiguous variants, tri- and multi-allelic variants and low imputation quality (imputation INFO score < 0.3). For the GBMI discovery datasets, leave-one-biobank-out meta-analysis using the inverse-variance weighted meta-analysis strategy was applied^18^.

Target datasets: We used 9 biobanks, i.e., BioBank Japan (BBJ)^50^, BioVU^51^, Lifelines^52^, UK Biobank (UKBB)^53^, Ontario Health Study (OHS)^54^, Estonian Biobank (ESTBB)^55^, FinnGen, Michigan Genomics Initiative (MGI)^56^ and Trøndelag Health Study (HUNT)^57^, as the target datasets, which were independent from the datasets included in the discovery GWAS. Brief descriptions about these biobanks can be found in Zhou et al.^18^. We removed individuals with genetic relatedness larger than 0.05 and applied the same filters as the discovery GWAS for SNPs. In addition, only common SNPs with MAF > 1% were retained.

### Genetic architecture of 14 endpoints in GBMI

SBayesS is a summary-level based method utilizing a Bayesian mixed linear model, which can report key parameters describing the genetic architecture of complex traits^20^. It only requires GWAS summary statistics and LD correlation matrix estimated from a reference panel. We ran SBayesS using the GWAS summary statistics from all 14 endpoints in GBMI, including meta-analyses on all ancestries and on EUR only in 19 biobanks^18^. We evaluated the SNP-based heritability 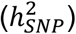, polygenicity (proportion of SNPs with nonzero effects) and the relationship between allele frequency and SNP effects (S). We used the shrunk LD matrix (i.e., a LD matrix ignoring small LD correlations due to sampling variance) on HapMap3 SNPs provided by GCTB software. The LD matrix was constructed based on 50K European individuals from UKBB. Note that we observed inflated SNP-based heritability estimates using effective sample size for each SNP and hence used the total GWAS sample size instead. We used other default settings in the software. We calculated the *p*-value of each parameter using Wald test to evaluate whether it was significantly different from 0. The 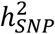 was further transformed into liability-scale with disease prevalence approximated as the case proportions in the GWAS summary statistics^58^.

### PRS construction

P+T: P+T is used to clump quasi-independent trait-associated loci within a LD window size using a specific LD *r*^2^ threshold. We first ran P+T in the UKBB and BBJ using a LD *r*^2^ threshold of 0.1 and a LD window (LD_win_) of 250Kb. We performed the analysis on both HapMap3 SNPs and genome-wide SNPs. We constructed PRS using *--score* implemented in Plink v1.9^59^ using 13 different *p*-value thresholds (5 × 10^−8^, 5 × 10^−7^, 1 × 10^−6^, 5 × 10^−6^, 5 × 10^−5^, 5 × 10^−4^, 5 × 10^−3^, 0.01, 0.05, 0.1, 0.2, 0.5, 1). We further explored how per-variant filtering based on effective sample sizes (*N*_*eff*_) and MAF thresholds would affect the prediction performance. We used three thresholds to retain variants by their *N*_*eff*_: >0%, >50%, and >80% of *N*_*eff*_ compared to the total ones and also three MAF filters: 0.01, 0.05 and 0.1. In the UKBB, we also explored the impact of optimizing LD parameters on prediction performance by using different combinations of LD_win_ (250, 500, 1000, and 2000Kb) and LD *r*^2^ thresholds (0.01, 0.02, 0.05, 0.1, 0.2, and 0.05) with the following flags: *--clump-p1 1 --clump-p2 1 --clump-r2* LD_win_ *--clump-kb r*^2^ in Plink v1.9. For each population in the specific biobank, we randomly split the individuals into two even parts. One part was used as a validation cohort to fine-tune the parameters and the other part was used as the test cohort to evaluate the performance of PRS. To explore the impact of tuning cohorts on target populations with diverse ancestries such as UKBB in this study, we also used 10,000 EUR samples, not included in the discovery GWAS and independent from the test cohort, as the tuning cohort.

PRS-CS: PRS-CS^27^ is a Bayesian regression framework which enables continuous shrinkage priors on SNP effects to infer their posterior mean effects. We ran PRS-CS using both the grid and auto models in the UKBB. In the grid model, we used a series of global shrinkage parameters (phi = 1 × 10^−6^, 1 × 10^−5^, 1 × 10^−4^, 1 × 10^−3^, 0.01, 0.1, 1), with lower phi values suggesting less polygenic genetic architecture and vice versa for more polygenic genetic architecture. For the auto model, PRS-CS will learn the phi parameter from the discovery GWAS without requiring post-hoc tuning. We used both total GWAS sample size and effective sample size as input for PRS-CS and found little difference, suggesting that PRS-CS is insensitive to the input of GWAS sample size. We hence used the effective sample size for subsequent analyses in this study. We used the default settings for other parameters. We generalized the auto model for all endpoints in both UKBB and BBJ. When comparing the two models, we selected the optimal phi parameter from the grid model based on the highest prediction accuracy in the target population.

### LD reference panel

Both P+T and PRS-CS are summary-level based PRS prediction methods, utilizing GWAS summary statistics and an LD reference panel. To explore the impact of LD reference panels on prediction performance, we used LD reference panels of different ancestral compositions, varying sample sizes and SNP density. Specifically, we used four global ancestry groups, i.e., European (EUR), South-Asian (SAS), East-Asian (EAS) and African (AFR), from 1000G Phase 3 (1KG) as LD reference panels for P+T. Further, we randomly sampled a subset of individuals with sample sizes of 500, 5000, 10,000 and 50,000 from UKBB-EUR to analyze how the sample sizes of LD reference panel would affect prediction accuracy for P+T. Moreover, we ran P+T on both the HapMap3 SNP set and a denser SNP set with genome-wide SNPs. We ran PRS-CS with the LD matrix provided by PRS-CS software^27^, which are based on both 1KG and UKBB populations from those four ancestry groups and Admixed American population (AMR). We performed those analyses using leave-UKBB-out GWAS in GBMI and evaluated the prediction performance in diverse ancestry groups in the UKBB.

To explore how well EUR-based LD reference approximated the LD of multi-ancestry GWAS in GBMI, we ran LD score regression (LDSC) to estimate the attenuation ratio statistic^21^. The values of attenuation ratio larger than 0.2 suggest a strong LD mismatch between GWAS summary statistics and LD reference panel. We performed LDSC analyses on different GWAS, including GBMI GWAS from meta-analyses on all ancestries, EUR only and leave-one-biobank-out.

### Evaluation of prediction performance

After constructing PRS, we evaluated the prediction performance in the independent target datasets. We used a logistic regression to calculate the Nagelkerke’s *R*^2^ and variance on the liability-scale explained by PRS as described previously^58^. Area under the receiver operating characteristic curve (AUC) was also reported for full models with additional covariates and models including PRS only. We used bootstrap with 1000 replicates to estimate their corresponding 95% confidence intervals (CIs). Note that the proportion of cases in each ancestry in the target dataset was approximated as the disease population prevalence. The same covariates (usually age, sex and 20 genotypic principal components, PCs) used in the GWAS analyses were included in the full regression model as phenotype ∼ PRS + covariates. We also calculated the average *R*^2^ on the liability scale and AUC across biobanks (denoted as 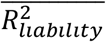 and 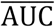, respectively) in each ancestry by weighting the effective sample size of each biobank for each endpoint. Further, we divided the target individuals into deciles based on the ranking of PRS distribution. We compared the odds ratio (OR) of the top decile relative to those ranked as the bottom, the middle and the remaining, when using the first decile as the referenced group. For endpoints presented in two or more biobanks, we calculated the averaged OR using the inverse variance weighted method and the coefficient of variation of OR (CoeffVar_OR_) as SD(OR)/mean(OR).

### Resource Availability

#### Data and Code Availability

The all-biobank and ancestry-specific GWAS summary statistics are publicly available for downloading at https://www.globalbiobankmeta.org/resources and browsed at the PheWeb Browser http://results.globalbiobankmeta.org/. The PRS weights re-estimated using PRC-CS-auto for multi-ancestry GWAS including all biobanks and leave-UKBB-out multi-ancestry GWAS have been uploaded to PGS Catalog (https://www.pgscatalog.org/) under the study ID PGP000262. 1000 Genome Phase 3 data can be accessed at ftp://ftp.1000genomes.ebi.ac.uk/vol1/ftp/data_collections/1000_genomes_project/data. We used UKB data via application 31063. The software used in this study can be found at: Plink (https://www.cog-genomics.org/plink/), PRS-CS (https://github.com/getian107/PRScs), SBayesS/GCTB (https://cnsgenomics.com/software/gctb/). The codes used in this study can be found in the github repository: https://github.com/globalbiobankmeta/PRS.

## Notes

### Summary of Updates

1.We have extended P+T analysis using different parameters (LD r2 threshold, LD window and p-value threshold) to all endpoints rather than only asthma in the previous manuscript. 2.We have added new results of P+T and revised the section Optimal prediction performance using heuristic methods depends on phenotype-specific genetic architecture. The main conclusion is largely consistent with previous analyses based on asthma. 3.We have substantially reorganized and revised the discussion section to clarify the impact and comprehensiveness of this study using GBMI resources, including the effects of multi-ancestry and heterogeneous GWAS discovery data, multiple endpoints with spanning genetic architecture as well as prevelances, and PRS methods on prediction performance in diverse target cohorts. 4.We have added a schematic figure (Figure S13) and a table (Table S5) to clarify recommendations and guidelines for PRS analyses using multi-ancestry GWAS.

## References

1. Abul-Husn NS, Kenny EE. Personalized Medicine and the Power of Electronic Health Records. Cell. 2019 Mar 21;177(1):58–69.

2. Inouye M, Abraham G, Nelson CP, Wood AM, Sweeting MJ, Dudbridge F, et al. Genomic Risk Prediction of Coronary Artery Disease in 480,000 Adults: Implications for Primary Prevention. J Am Coll Cardiol. 2018 Oct 16;72(16):1883–93.

3. Torkamani A, Wineinger NE, Topol EJ. The personal and clinical utility of polygenic risk scores. Nat Rev Genet. 2018 Sep;19(9):581–90.

4. Lewis CM, Vassos E. Polygenic risk scores: from research tools to clinical instruments. Genome Med. 2020 May 18;12(1):44.

5. Mavaddat N, Michailidou K, Dennis J, Lush M, Fachal L, Lee A, et al. Polygenic Risk Scores for Prediction of Breast Cancer and Breast Cancer Subtypes. Am J Hum Genet. 2019 Jan 3;104(1):21–34.

6. Landi I, Kaji DA, Cotter L, Van Vleck T, Belbin G, Preuss M, et al. Prognostic value of polygenic risk scores for adults with psychosis. Nat Med. 2021 Sep 6;1–6.

7. Dudbridge F, Pashayan N, Yang J. Predictive accuracy of combined genetic and environmental risk scores. Genet Epidemiol. 2018 Feb;42(1):4–19.

8. Craig JE, Han X, Qassim A, Hassall M, Cooke Bailey JN, Kinzy TG, et al. Multitrait analysis of glaucoma identifies new risk loci and enables polygenic prediction of disease susceptibility and progression. Nat Genet. 2020 Jan 20;52(2):160–6.

9. Ni G, Zeng J, Revez JA, Wang Y, Zheng Z, Ge T, et al. A Comparison of Ten Polygenic Score Methods for Psychiatric Disorders Applied Across Multiple Cohorts. Biol Psychiatry. 2021 Nov 1;90(9):611–20.

10. Ma Y, Zhou X. Genetic prediction of complex traits with polygenic scores: a statistical review. Trends Genet. 2021 Nov;37(11):995–1011.

11. Kulm S, Marderstein A, Mezey J. A systematic framework for assessing the clinical impact of polygenic risk scores [Internet]. MedRxiv. 2021. Available from: https://www.medrxiv.org/content/10.1101/2020.04.06.20055574v2.full-text

12. Majara L, Kalungi A, Koen N, Zar H, Stein DJ, Kinyanda E, et al. Low generalizability of polygenic scores in African populations due to genetic and environmental diversity [Internet]. Cold Spring Harbor Laboratory. 2021 [cited 2021 Jan 28]. p. 2021.01.12.426453. Available from: https://www.biorxiv.org/content/10.1101/2021.01.12.426453v1.abstract

13. Martin AR, Kanai M, Kamatani Y, Okada Y, Neale BM, Daly MJ. Clinical use of current polygenic risk scores may exacerbate health disparities. Nat Genet. 2019 Apr;51(4):584–91.

14. Mostafavi H, Harpak A, Agarwal I, Conley D, Pritchard JK, Przeworski M. Variable prediction accuracy of polygenic scores within an ancestry group. Elife. 2020 Jan 30;9:e48376.

15. Martin AR, Daly MJ, Robinson EB, Hyman SE, Neale BM. Predicting Polygenic Risk of Psychiatric Disorders. Biol Psychiatry. 2019 Jul 15;86(2):97–109.

16. Ruan Y, Anne Feng YC, Chen CY, Lam M, Sawa A, Martin AR, et al. Improving polygenic prediction in ancestrally diverse populations [Internet]. medRxiv. 2021. Available from: http://medrxiv.org/lookup/doi/10.1101/2020.12.27.20248738

17. Weissbrod O, Kanai M, Shi H, Gazal S, Peyrot W, Khera A, et al. Leveraging fine-mapping and non-European training data to improve trans-ethnic polygenic risk scores [Internet]. medRxiv. 2021. Available from: http://medrxiv.org/lookup/doi/10.1101/2021.01.19.21249483

18. Zhou W, Kanai M, Wu KHH, Humaira R, Tsuo K, Hirbo JB, et al. Global Biobank Meta-analysis Initiative: powering genetic discovery across human diseases [Internet]. medRxiv. 2021. Available from: http://medrxiv.org/lookup/doi/10.1101/2021.11.19.21266436

19. Zhou W, Kanai M, Wu KHH, Humaira R, Tsuo K, Hirbo JB, et al. Global Biobank Meta-analysis Initiative: powering genetic discovery across human diseases [Internet]. MedRxiv. 2021. Available from: http://medrxiv.org/lookup/doi/10.1101/2021.11.19.21266436

20. Zeng J, Xue A, Jiang L, Lloyd-Jones LR, Wu Y, Wang H, et al. Widespread signatures of natural selection across human complex traits and functional genomic categories. Nat Commun. 2021 Feb 19;12(1):1164.

21. Bulik-Sullivan BK, Loh PR, Finucane HK, Ripke S, Yang J, Schizophrenia Working Group of the Psychiatric Genomics Consortium, et al. LD Score regression distinguishes confounding from polygenicity in genome-wide association studies. Nat Genet. 2015 Mar;47(3):291–5.

22. Yengo L, Vedantam S, Marouli E, Sidorenko J, Bartell E, Sakaue S, et al. A Saturated Map of Common Genetic Variants Associated with Human Height from 5.4 Million Individuals of Diverse Ancestries [Internet]. bioRxiv. 2022 [cited 2022 Jan 11]. p. 2022.01.07.475305. Available from: https://www.biorxiv.org/content/10.1101/2022.01.07.475305v1?rss=1

23. O’Connor LJ, Schoech AP, Hormozdiari F, Gazal S, Patterson N, Price AL. Extreme Polygenicity of Complex Traits Is Explained by Negative Selection. Am J Hum Genet. 2019 Sep 5;105(3):456–76.

24. Zhang Y, Qi G, Park JH, Chatterjee N. Estimation of complex effect-size distributions using summary-level statistics from genome-wide association studies across 32 complex traits. Nat Genet. 2018 Sep;50(9):1318–26.

25. Ware EB, Schmitz LL, Faul J, Gard A, Mitchell C, Smith JA, et al. Heterogeneity in polygenic scores for common human traits [Internet]. bioRxiv. 2017. p. 106062. Available from: https://www.biorxiv.org/content/10.1101/106062v1

26. Choi SW, Mak TSH, O’Reilly PF. Tutorial: a guide to performing polygenic risk score analyses. Nat Protoc [Internet]. 2020 Jul 24; Available from: http://dx.doi.org/10.1038/s41596-020-0353-1

27. Ge T, Chen CY, Ni Y, Feng YCA, Smoller JW. Polygenic prediction via Bayesian regression and continuous shrinkage priors. Nat Commun. 2019 Apr 16;10(1):1776.

28. Martin AR, Gignoux CR, Walters RK, Wojcik GL, Neale BM, Gravel S, et al. Human Demographic History Impacts Genetic Risk Prediction across Diverse Populations. Am J Hum Genet. 2017 Apr 6;100(4):635–49.

29. Duncan L, Shen H, Gelaye B, Meijsen J, Ressler K, Feldman M, et al. Analysis of polygenic risk score usage and performance in diverse human populations. Nat Commun. 2019 Jul 25;10(1):1–9.

30. Wang Y, Guo J, Ni G, Yang J, Visscher PM, Yengo L. Theoretical and empirical quantification of the accuracy of polygenic scores in ancestry divergent populations. Nat Commun. 2020 Jul 31;11(1):3865.

31. Borish L, Culp JA. Asthma: a syndrome composed of heterogeneous diseases. Ann Allergy Asthma Immunol. 2008 Jul;101(1):1–8; quiz 8–11, 50.

32. Lo Faro, Bhattacharya, Zhou, Zhou, Wang Läll, et al. Global Biobank Meta-Analysis Initiative: A genome-wide association meta-analysis identifies novel primary open-angle glaucoma loci and shared biology with vascular mechanisms and cell proliferation. In preparation. 2021;

33. Faro VL, Bhattacharya A, Zhou W, Zhou D, Wang Y, Läll K, et al. Genome-wide association meta-analysis identifies novel ancestry-specific primary open-angle glaucoma loci and shared biology with vascular mechanisms and cell proliferation [Internet]. medRxiv. 2021. Available from: https://www.medrxiv.org/content/10.1101/2021.12.16.21267891.abstract

34. Surakka I, Wu KH, Hornsby W, Wolford BN, Shen F, Zhou W, et al. Multi-ancestry meta-analysis identifies 2 novel loci associated with ischemic stroke and reveals heterogeneity of effects between sexes and ancestries [Internet]. bioRxiv. 2022. Available from: https://www.medrxiv.org/content/10.1101/2022.02.28.22271647.abstract

35. Partanen JJ, Häppölä P, Zhou W, Lehisto AA, Ainola M, Sutinen E, et al. Leveraging global multi-ancestry meta-analysis in the study of Idiopathic Pulmonary Fibrosis genetics [Internet]. bioRxiv. 2021. Available from: https://www.medrxiv.org/content/10.1101/2021.12.29.21268310.abstract

36. Wand H, Lambert SA, Tamburro C, Iacocca MA, O’Sullivan JW, Sillari C, et al. Improving reporting standards for polygenic scores in risk prediction studies. Nature. 2021 Mar;591(7849):211–9.

37. Privé F, Aschard H, Carmi S, Folkersen L, Hoggart C, O’Reilly PF, et al. Portability of 245 polygenic scores when derived from the UK Biobank and applied to 9 ancestry groups from the same cohort. Am J Hum Genet. 2022 Feb 3;109(2):373.

38. Wang Y, Tsuo K, Kanai M, Neale BM, Martin AR. Challenges and Opportunities for Developing More Generalizable Polygenic Risk Scores. Annu Rev Biomed Data Sci [Internet]. 2022 May 16; Available from: http://dx.doi.org/10.1146/annurev-biodatasci-111721-074830

39. Graham SE, Clarke SL, Wu KH, Lin K, Millwood IY, Mahajan A, et al. The power of genetic diversity in genome-wide association studies of lipids. Nature [Internet]. 2021 [cited 2021 Dec 10]; Available from: https://ora.ox.ac.uk/objects/uuid:5d0c9801-0dbf-4d5d-8d19-95606c30a2c0

40. Mak TSH, Porsch RM, Choi SW, Zhou X, Sham PC. Polygenic scores via penalized regression on summary statistics. Genet Epidemiol. 2017 Sep;41(6):469–80.

41. Miao J, Guo H, Song G, Zhao Z, Hou L, Lu Q. Quantifying portable genetic effects and improving cross-ancestry genetic prediction with GWAS summary statistics [Internet]. bioRxiv. 2022 [cited 2022 Jun 17]. p. 2022.05.26.493528. Available from: https://www.biorxiv.org/content/10.1101/2022.05.26.493528v1

42. Blatt M, Gusev A, Polyakov Y, Goldwasser S. Secure large-scale genome-wide association studies using homomorphic encryption. Proc Natl Acad Sci U S A. 2020 May 26;117(21):11608–13.

43. Márquez-Luna C, Loh PR, South Asian Type 2 Diabetes (SAT2D) Consortium, SIGMA Type 2 Diabetes Consortium, Price AL. Multiethnic polygenic risk scores improve risk prediction in diverse populations. Genet Epidemiol. 2017 Dec;41(8):811–23.

44. Tsuo, Zhou, Wang, Kanai, Namba, Gupta, et al. Multi-ancestry meta-analysis of asthma identifies novel associations and highlights shared genetic architecture across biobanks and traits. In preparation. 2021;

45. Meisner A, Kundu P, Chatterjee N. Case-Only Analysis of Gene-Environment Interactions Using Polygenic Risk Scores. Am J Epidemiol. 2019 Nov 1;188(11):2013–20.

46. Loika Y, Irincheeva I, Culminskaya I, Nazarian A, Kulminski AM. Polygenic risk scores: pleiotropy and the effect of environment. Geroscience. 2020 Dec;42(6):1635–47.

47. Atkinson EG, Maihofer AX, Kanai M, Martin AR, Karczewski KJ, Santoro ML, et al. Tractor uses local ancestry to enable the inclusion of admixed individuals in GWAS and to boost power. Nat Genet. 2021 Feb;53(2):195–204.

48. Cavazos TB, Witte JS. Inclusion of variants discovered from diverse populations improves polygenic risk score transferability. Human Genetics and Genomics Advances. 2021 Jan 14;2(1):100017.

49. Marnetto D, Pärna K, Läll K, Molinaro L, Montinaro F, Haller T, et al. Ancestry deconvolution and partial polygenic score can improve susceptibility predictions in recently admixed individuals. Nat Commun. 2020 Apr 2;11(1):1–9.

50. Nagai A, Hirata M, Kamatani Y, Muto K, Matsuda K, Kiyohara Y, et al. Overview of the BioBank Japan Project: Study design and profile. J Epidemiol. 2017 Mar;27(3S):S2–8.

51. Bowton EA, Collier SP, Wang X, Sutcliffe CB, Van Driest SL, Couch LJ, et al. Phenotype-Driven Plasma Biobanking Strategies and Methods. J Pers Med. 2015 May 14;5(2):140–52.

52. Scholtens S, Smidt N, Swertz MA, Bakker SJL, Dotinga A, Vonk JM, et al. Cohort Profile: LifeLines, a three-generation cohort study and biobank. Int J Epidemiol. 2015 Aug;44(4):1172–80.

53. Bycroft C, Freeman C, Petkova D, Band G, Elliott LT, Sharp K, et al. The UK Biobank resource with deep phenotyping and genomic data. Nature. 2018 Oct 10;562(7726):203–9.

54. Dummer TJB, Awadalla P, Boileau C, Craig C, Fortier I, Goel V, et al. The Canadian Partnership for Tomorrow Project: a pan-Canadian platform for research on chronic disease prevention. CMAJ. 2018 Jun 11;190(23):E710–7.

55. Leitsalu L, Haller T, Esko T, Tammesoo ML, Alavere H, Snieder H, et al. Cohort Profile: Estonian Biobank of the Estonian Genome Center, University of Tartu. Int J Epidemiol. 2015 Aug;44(4):1137–47.

56. Zawistowski, Fritsche, Pandit, Vanderwerff, Patil, Schmidt, et al. The Michigan Genomics Initiative: a biobank linking genotypes and electronic clinical records in Michigan Medicine patients. In preparation. 2021;

57. Krokstad S, Langhammer A, Hveem K, Holmen TL, Midthjell K, Stene TR, et al. Cohort Profile: the HUNT Study, Norway. Int J Epidemiol. 2013 Aug;42(4):968–77.

58. Lee SH, Goddard ME, Wray NR, Visscher PM. A better coefficient of determination for genetic profile analysis. Genet Epidemiol. 2012 Apr;36(3):214–24.

59. Chang CC, Chow CC, Tellier LC, Vattikuti S, Purcell SM, Lee JJ. Second-generation PLINK: rising to the challenge of larger and richer datasets. Gigascience. 2015 Feb 25;4:7.

