## Supplementary Figures and Tables for "Global biobank analyses provide lessons for developing polygenic risk scores across diverse cohorts"

**A**

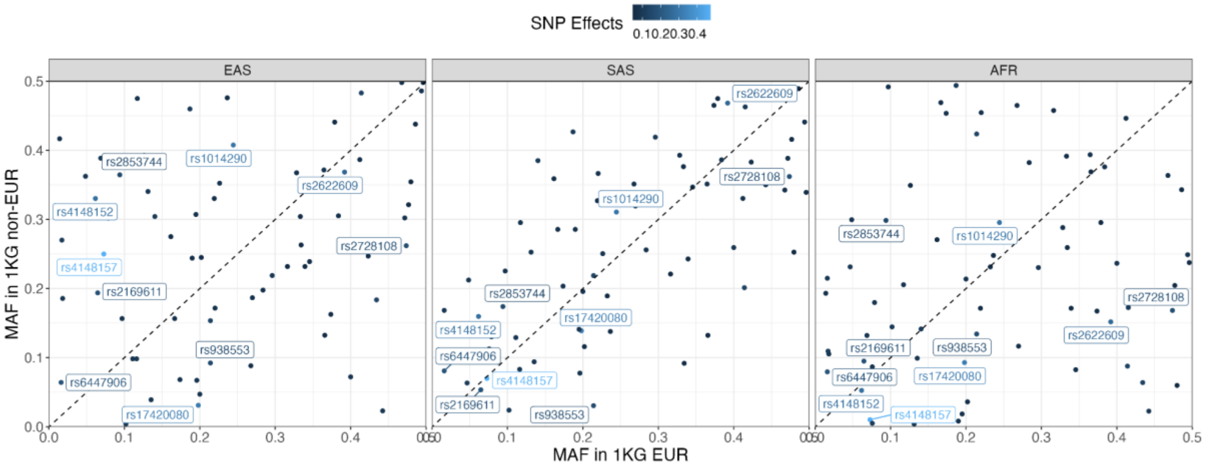

**B**

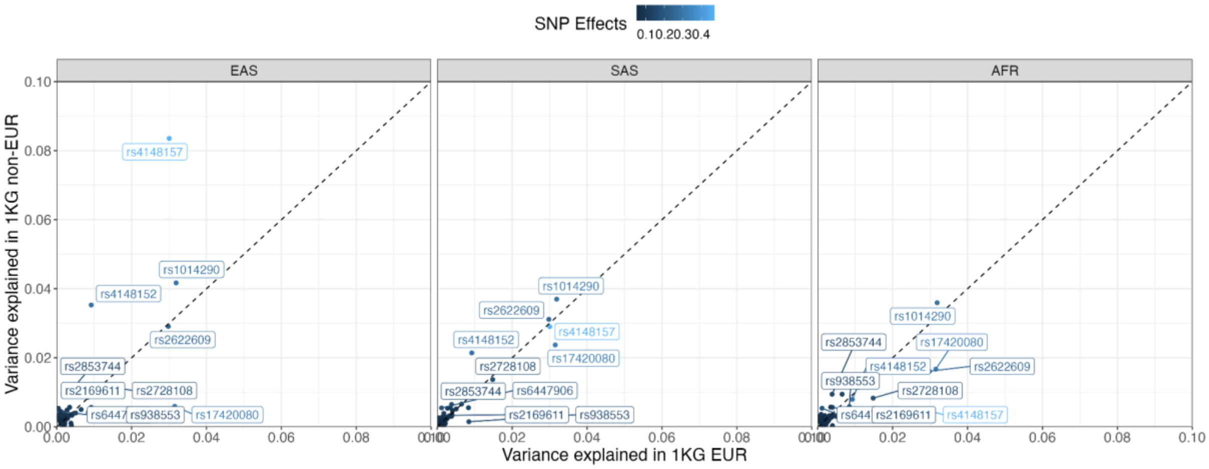

**Figure S1. Minor allele frequency and variance explained for gout-associated genome-wide significant SNPs in 1KG.**

We showed **A)** The Minor allele frequency distribution and **B**) variance explained for gout-associated GWS SNPs. The genome-wide significant SNPs were identified using P+T (see STAR Methods). The dashed line denotes y = x. The labeled SNPs have top 10 ranked SNP effects. Abbreviations: Europeans (EUR), South Asians (SAS), East Asians (EAS) and Africans (AFR).

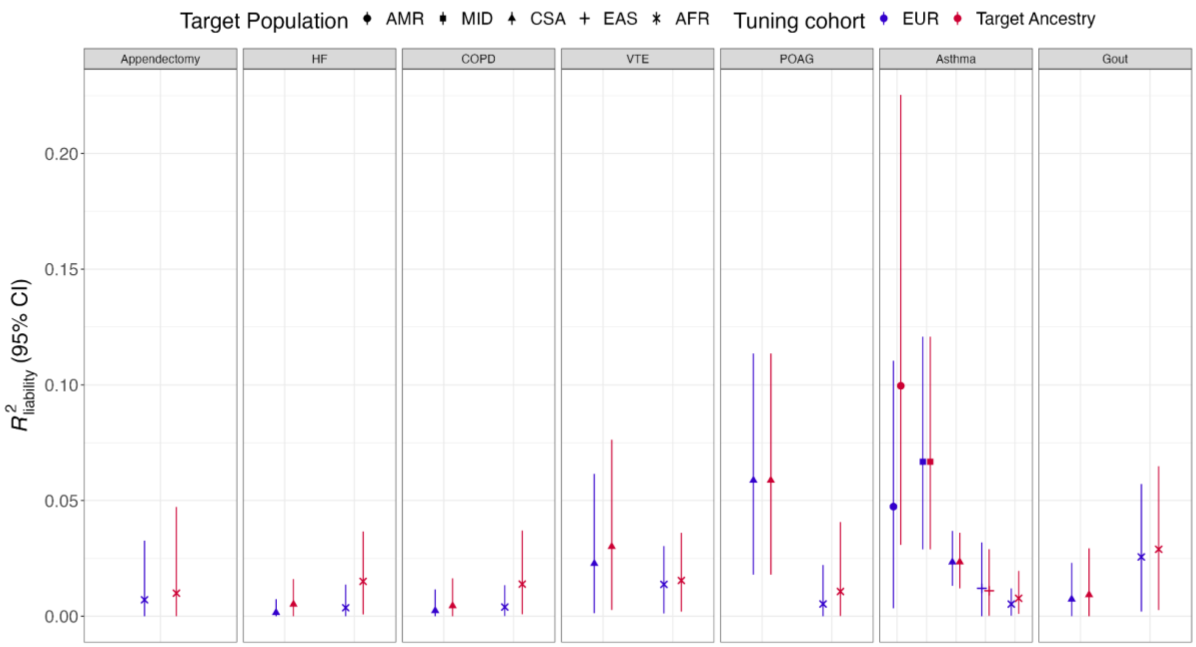

**Figure S2. The impact of tuning cohorts on the prediction performance of P+T.**

We evaluated the accuracy in the non-EUR populations of UKBB using different tuning cohorts. Tuning cohort labeled as “EUR” indicated that 10,000 EUR from UKBB was used as tuning cohort whilst the label “Target Ancestry” suggested we randomly split the target cohort into two equally distributed datasets, with one used as the tuning cohort (see details in STAR Methods). The phenotypes in columns were ranked based on the SNP-based heritability estimates using all ancestries (**see Figure 2**). Abbreviations: UK Biobank (UKBB), Biobank Japan (BBJ), Europeans (EUR), East Asians (EAS), chronic obstructive pulmonary disease (COPD), heart failure (HF), acute appendicitis (AcApp), venous thromboembolism (VTE), primary open-angle glaucoma (POAG), uterine cancer (UtC), abdominal aortic aneurysm (AAA), idiopathic pulmonary fibrosis (IPF), thyroid cancer (ThC).

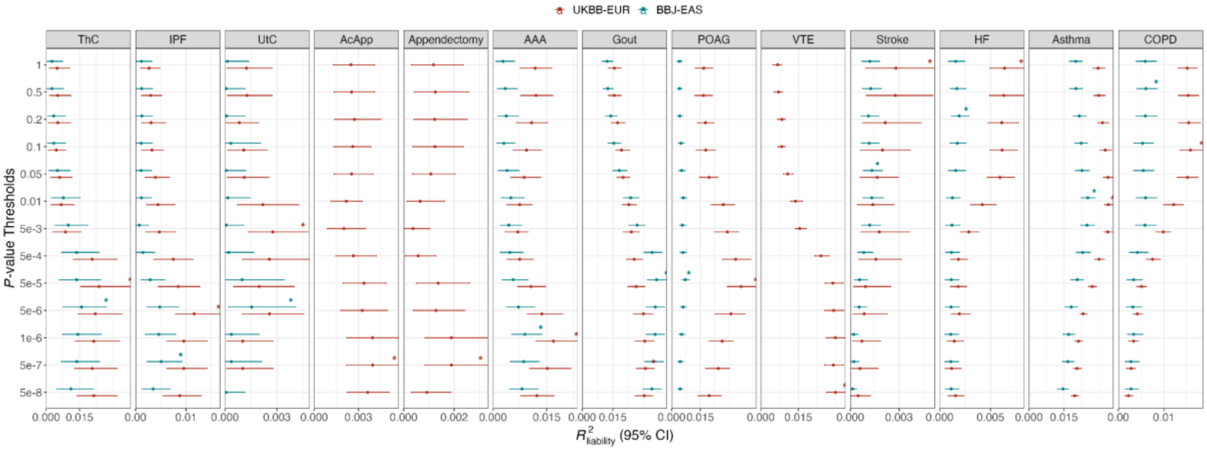

**Figure S3. The prediction performance of P+T using different *p*-value thresholds.**

We evaluated the accuracies in both UKBB-EUR and BBJ-EAS in the tuning cohort to select the optimal *p*-value thresholds. The asterisks indicate the optimal *p*-value threshold in each endpoint. The phenotypes in columns were ranked based on the polygenicity estimates using all ancestries (**see Figure 2**). Abbreviations: UK Biobank (UKBB), Boibank Japan (BBJ), Europeans (EUR), East Asians (EAS), chronic obstructive pulmonary disease (COPD), heart failure (HF), acute appendicitis (AcApp), venous thromboembolism (VTE), primary open-angle glaucoma (POAG), uterine cancer (UtC), abdominal aortic aneurysm (AAA), idiopathic pulmonary fibrosis (IPF), thyroid cancer (ThC).

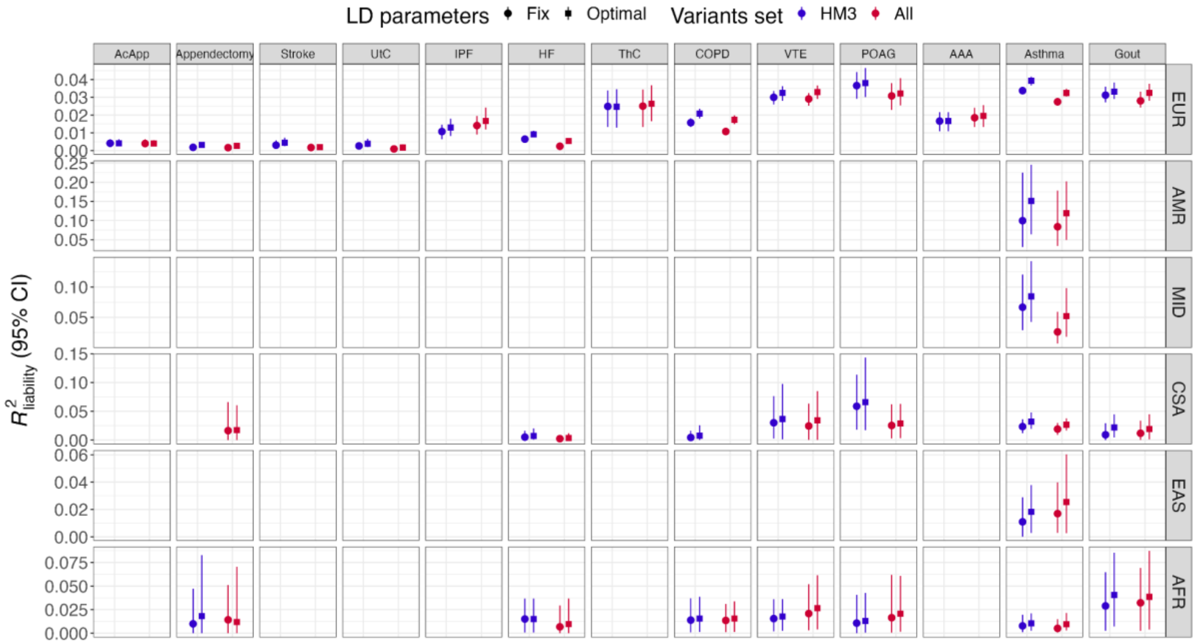

**Figure S4. Prediction performance of P+T in the UKBB using different optimization.**

We ran P+T with different combinations of *p*-value thresholds ( $5\times 10^{-8}$, $5\times 10^{-7}$, $1\times 10^{-6}$, $5\times 10^{-6}$, $5\times 10^{-5}$, $5\times 10^{-4}$, $5\times 10^{-3}$, 0.01, 0.05, 0.1, 0.2, 0.5 and 1), LD $r^2$ thresholds ($r^2$=0.01, 0.02, 0.05, 0.1, 0.2, and 0.5) and LD windows ($LD_{win}$=250, 500, 1000, and 2000Kb) for 13 endpoints in the UKBB (see STAR Methods). Both HapMap3 variants (HM3) and a denser genome-wide variant set (All) were analyzed.1KG-EUR was used as the LD reference panel in all analyses. The results using fixed LD parameters ($LD_{win}$=250, LD $r^2$=0.1) but optimizing *p*-value thresholds were reported as “Fix”, while the results of “Optimal” were based on all parameters optimization. We randomly split the target cohort into two equally distributed datasets, with one used as the tuning cohort to fine-tune hyper-parameters and the other used as test cohort to estimate prediction accuracy. Abbreviations: Europeans (EUR), Admixed Americans (AMR), Middle Eastern (MID), Central and South Asians (CSA), East Asians (EAS), Africans (AFR), chronic obstructive pulmonary disease (COPD), heart failure (HF), acute appendicitis (AcApp), venous thromboembolism (VTE), primary open-angle glaucoma (POAG), uterine cancer (UtC), abdominal aortic aneurysm (AAA), idiopathic pulmonary fibrosis (IPF), thyroid cancer (ThC).

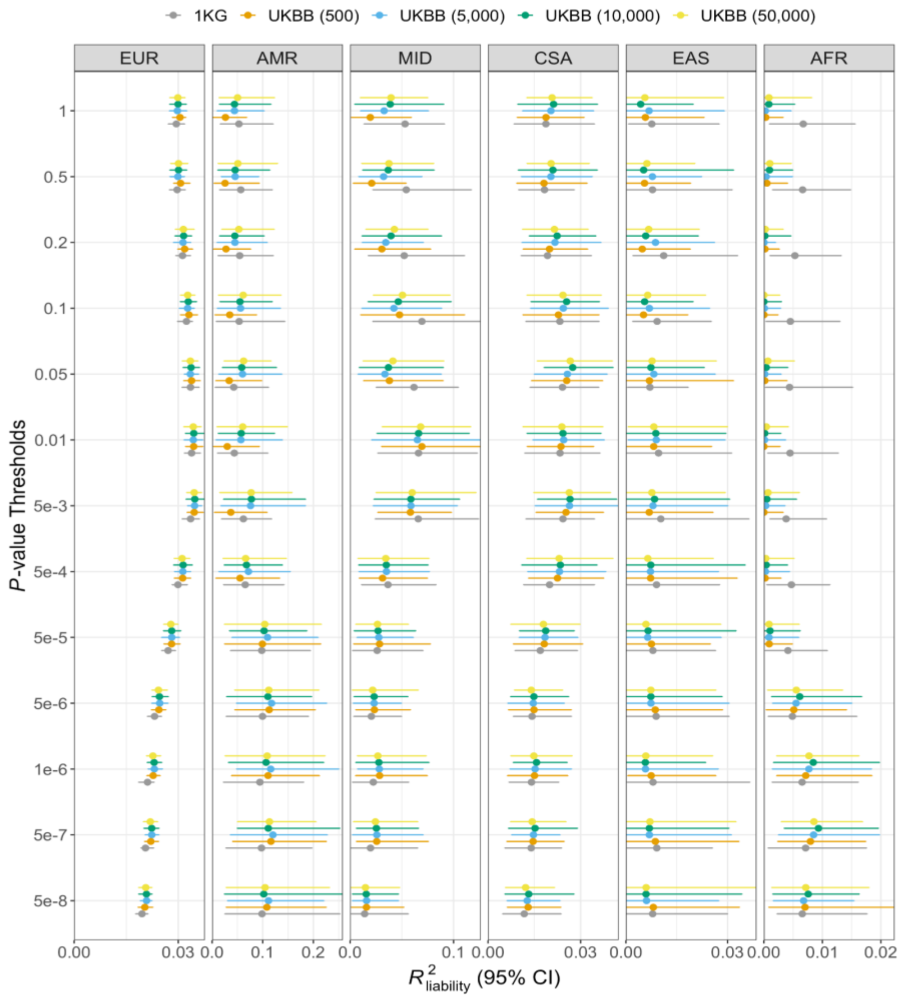

**Figure S5. The impact of LD reference panels' sample sizes on P+T prediction performance.**

We varied the sample sizes of EUR-based LD reference panels from 500 to 50,000. The accuracies were evaluated for asthma in the UKBB. We randomly split the target cohort into two equally distributed datasets, with one used as the tuning cohort to fine-tune hyper-parameters and the other used as test cohort to estimate prediction accuracy. Abbreviations: Europeans (EUR), Admixed Americans (AMR), Middle Eastern (MID), Central and South Asians (CSA), East Asians (EAS), Africans (AFR).

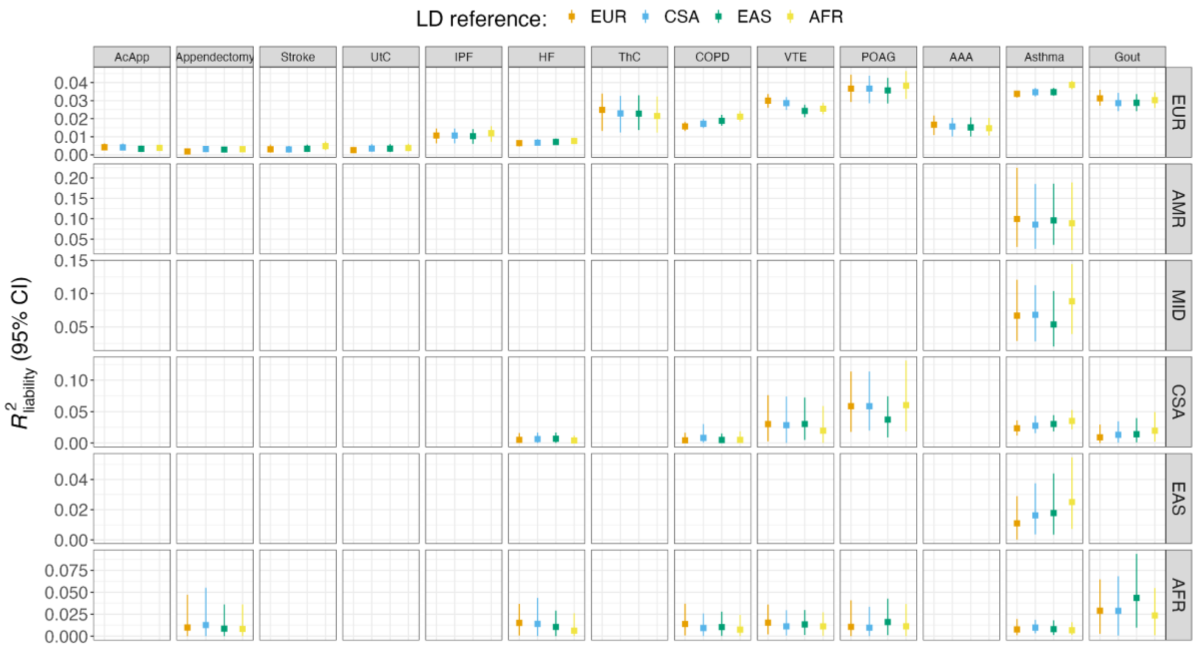

**Figure S6. The impact of LD reference panels' ancestral composition on P+T performance.**

We used different ancestral populations from 1KG as LD reference to run P+T on HapMap3 SNPs using fixed LD parameters. We randomly split each target population in the UKBB into two equally distributed datasets, with one used as the tuning cohort to fine-tune hyper-parameters and the other used as test cohort to estimate prediction accuracy. We found there was no significant difference of accuracies using different ancestral LD reference panels. Abbreviations: Europeans (EUR), Admixed Americans (AMR), Middle Eastern (MID), Central and South Asians (CSA), East Asians (EAS), Africans (AFR), chronic obstructive pulmonary disease (COPD), heart failure (HF), acute appendicitis (AcApp), venous thromboembolism (VTE), primary open-angle glaucoma (POAG), uterine cancer (UtC), abdominal aortic aneurysm (AAA), idiopathic pulmonary fibrosis (IPF), thyroid cancer (ThC).

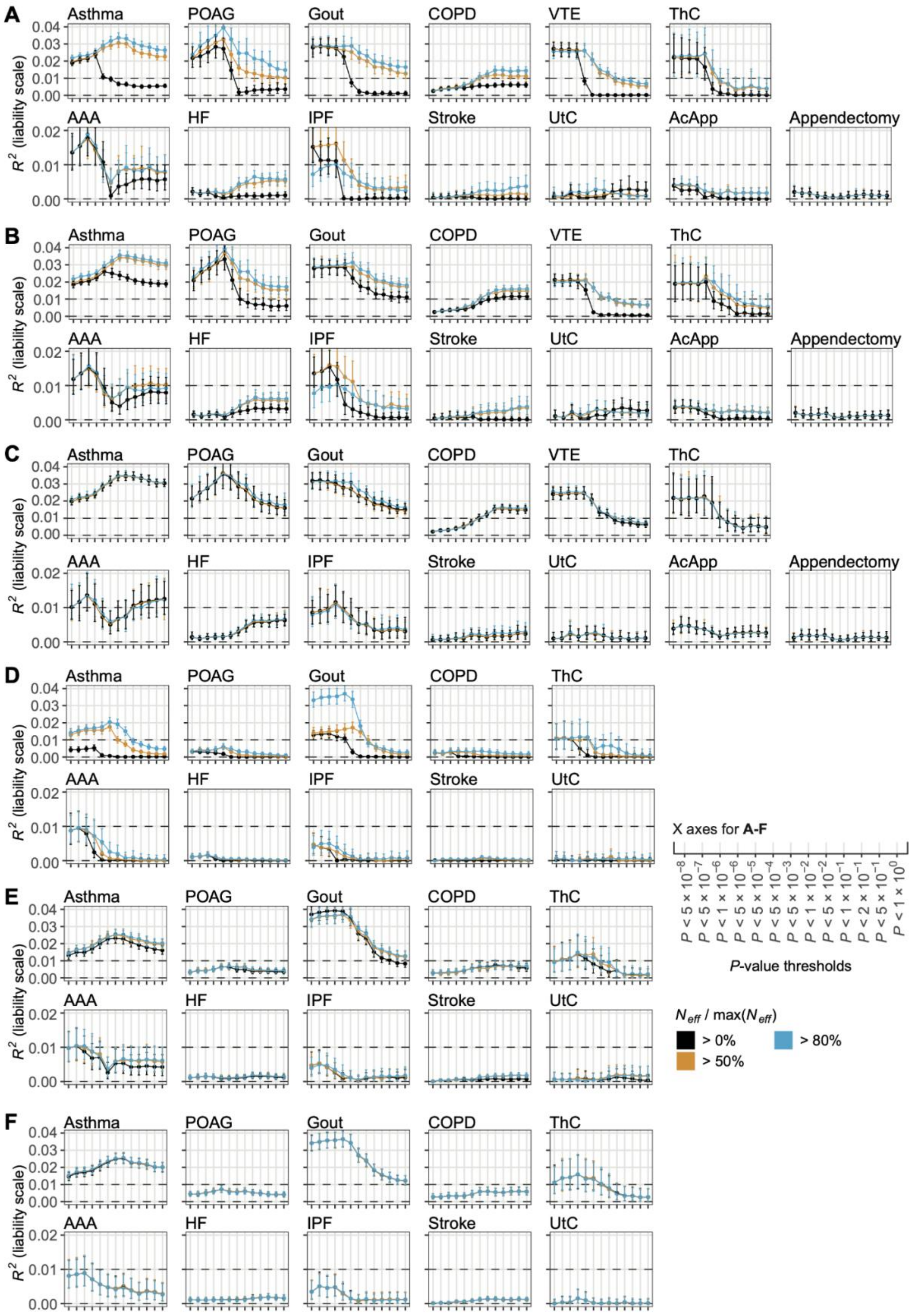
A
B
C
D
E
F
Asthma
POAG
Gout
COPD
VTE
ThC
AAA
HF
IPF
Stroke
UtC
AcApp
Appendectomy
$R^2$ (liability scale)
X axes for **A-F**
$P < 5 \times 10^{-8}$
$P < 5 \times 10^{-7}$
$P < 1 \times 10^{-6}$
$P < 5 \times 10^{-6}$
$P < 5 \times 10^{-5}$
$P < 5 \times 10^{-4}$
$P < 5 \times 10^{-3}$
$P < 1 \times 10^{-2}$
$P < 5 \times 10^{-2}$
$P < 1 \times 10^{-1}$
$P < 2 \times 10^{-1}$
$P < 5 \times 10^{-1}$
$P < 1 \times 10^{0}$
*P*-value thresholds
$N_{eff}$ / max($N_{eff}$)
> 0%
> 50%
> 80%

**Figure S7. PRS predictive performance for P+T stratified by sample size heterogeneity.**

The $R^2$ of P+T for 13 endpoints for European samples in the UK Biobank (UKBB) (**A**-**C**) and East Asian samples in Biobank Japan (BBJ) (**D**-**F**). Clinical information for VTE, AcApp, and Appendectomy was not collected in BBJ. **A** and **D** show the $R^2$ of PRS without filtering by minor allele frequency (MAF), while the variants with MAF less than 0.1 were excluded for PRS calculation in **B** and **E**. The HapMap3 variants were used for PRS calculation in **C** and **F**. The full results showing the effect of per-variant effective sample size ($N_{eff}$) and minor allele frequency (MAF) filtering are shown in **Table S4**. Abbreviations: chronic obstructive pulmonary disease (COPD), heart failure (HF), acute appendicitis (AcApp), venous thromboembolism (VTE), primary open-angle glaucoma (POAG), uterine cancer (UtC), abdominal aortic aneurysm (AAA), idiopathic pulmonary fibrosis (IPF), thyroid cancer (ThC).

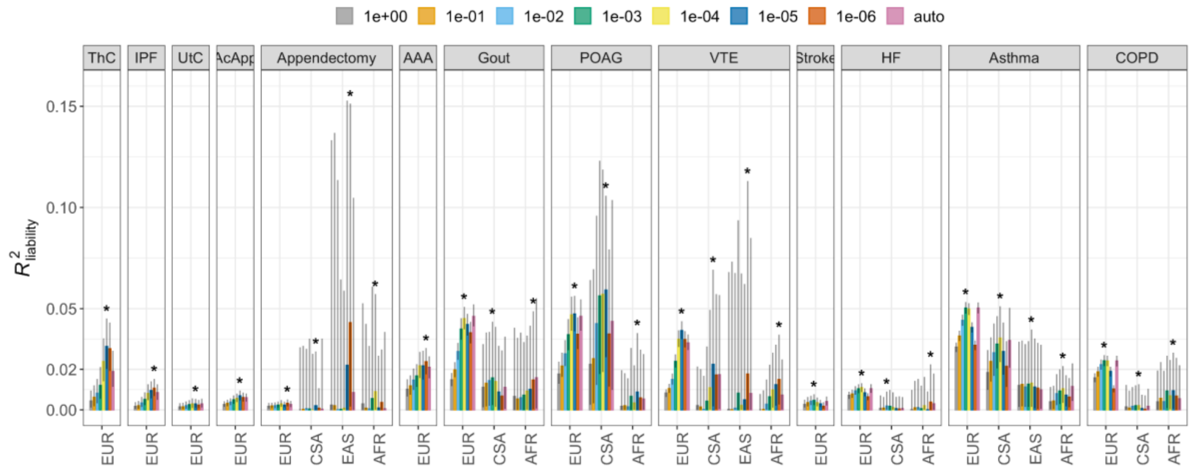

**Figure S8. Prediction performance of different PRS-CS models.**

We ran PRS-CS using both grid model and auto model (see STAR Methods). The asterisks indicate the optimized phi parameter with highest prediction accuracy achieved by grid model in each target ancestry in the UKBB. The phenotypes were ranked by the polygenicity using all ancestries as shown in **Figure 2**. Note that we removed the estimates in AMR and MID due to limited information as a result of small sample sizes. Abbreviations: Europeans (EUR), Admixed Americans (AMR), Middle Eastern (MID), Central and South Asians (CSA), East Asians (EAS), Africans (AFR), chronic obstructive pulmonary disease (COPD), heart failure (HF), acute appendicitis (AcApp), venous thromboembolism (VTE), primary open-angle glaucoma (POAG), uterine cancer (UtC), abdominal aortic aneurysm (AAA), idiopathic pulmonary fibrosis (IPF), thyroid cancer (ThC).

A

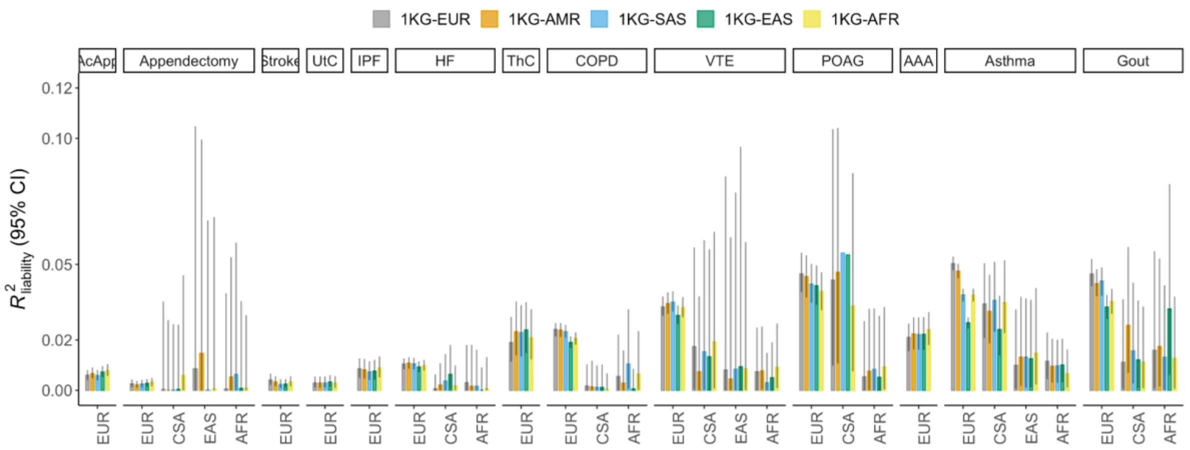

B

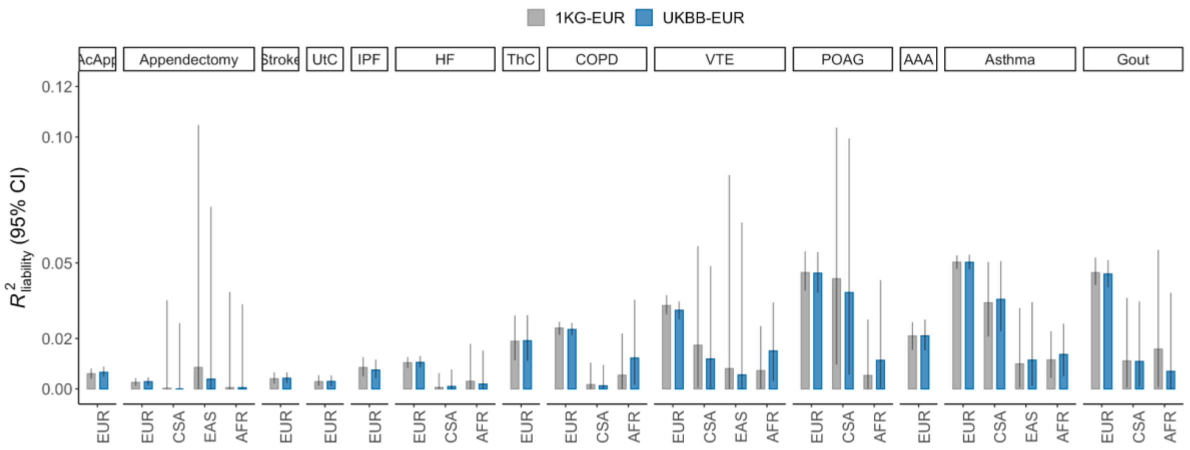

**Figure S9. The impact of LD reference panels on prediction accuracy using PRS-CS auto models.**

A) We used LD references from diverse ancestral populations in 1KG for running PRS-CS auto models. B) We used EUR LD reference from both 1KG and UKBB with different sample sizes. The phenotypes were ranked by the SNP-based heritability using all ancestries as shown in Figure 2. Note that we removed the estimates in AMR and MID due to limited information as a result of small sample sizes. Abbreviations: Europeans (EUR), Admixed Americans (AMR), Middle Eastern (MID), Central and South Asians (CSA), East Asians (EAS), Africans (AFR), chronic obstructive pulmonary disease (COPD), heart failure (HF), acute appendicitis (AcApp), venous thromboembolism (VTE), primary open-angle glaucoma (POAG), uterine cancer (UtC), abdominal aortic aneurysm (AAA), idiopathic pulmonary fibrosis (IPF), thyroid cancer (ThC).

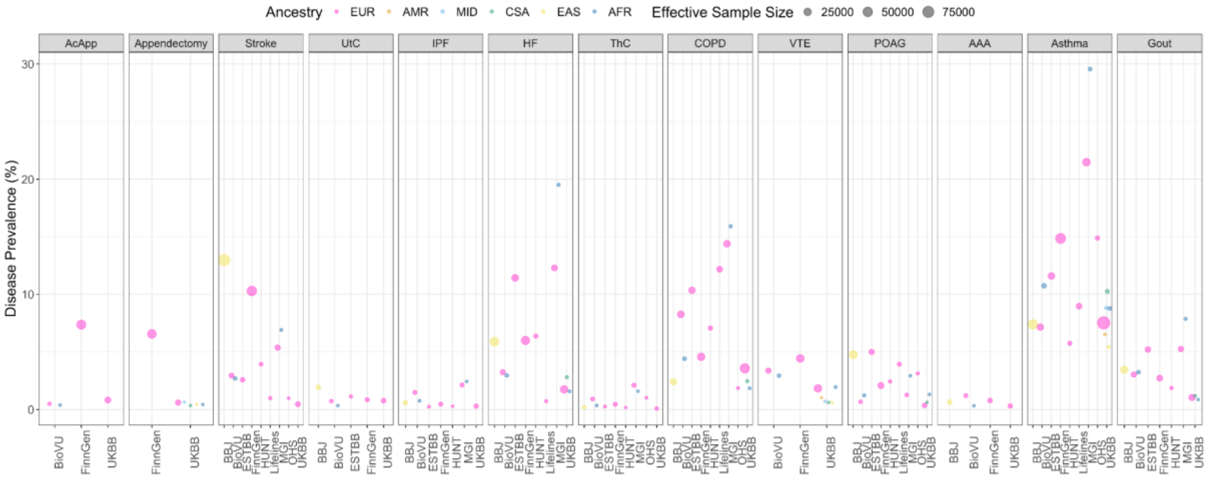

**Figure S10. The population prevalence and effective sample size of endpoints in GBMI for each biobank.**

Abbreviations: Europeans (EUR), Admixed Americans (AMR), Middle Eastern (MID), Central and South Asians (CSA), East Asians (EAS), Africans (AFR), chronic obstructive pulmonary disease (COPD), heart failure (HF), acute appendicitis (AcApp), venous thromboembolism (VTE), primary open-angle glaucoma (POAG), uterine cancer (UtC), abdominal aortic aneurysm (AAA), idiopathic pulmonary fibrosis (IPF), thyroid cancer (ThC).

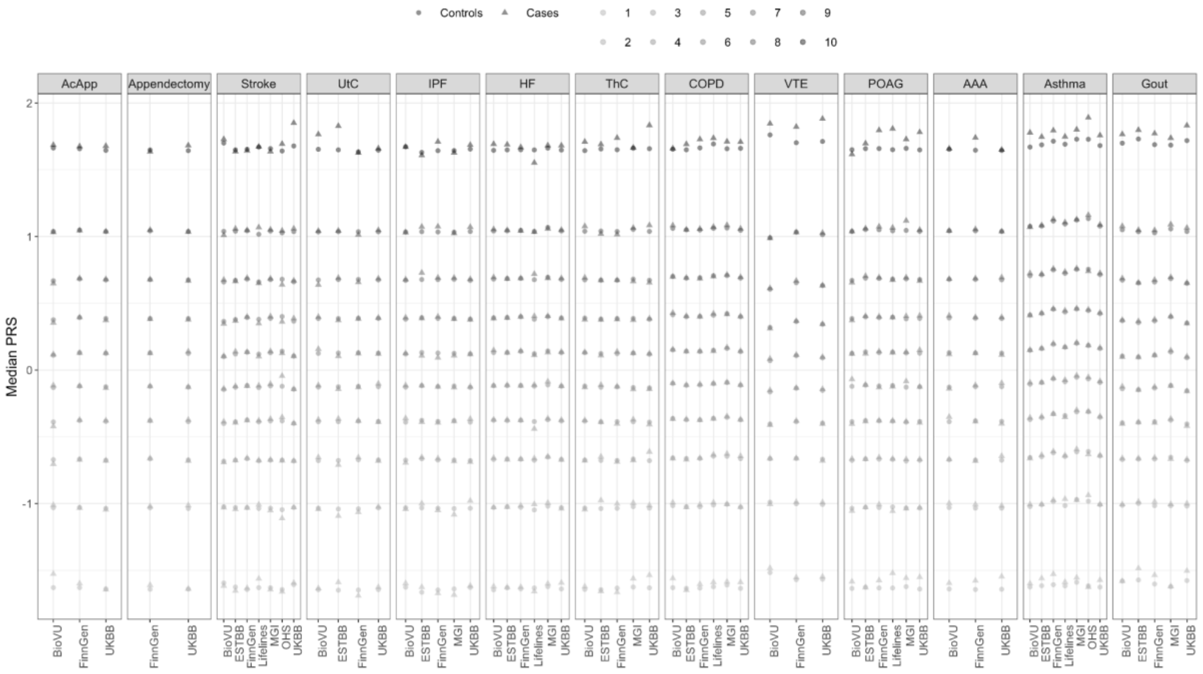

**Figure S11. The distribution of median PRS across biobanks in EUR.**

PRS was splitted into deciles while PRS in controls were normalized with mean of 0 and variance of 1. Abbreviations: Europeans (EUR), Admixed Americans (AMR), Middle Eastern (MID), Central and South Asians (CSA), East Asians (EAS), Africans (AFR), chronic obstructive pulmonary disease (COPD), heart failure (HF), acute appendicitis (AcApp), venous thromboembolism (VTE), primary open-angle glaucoma (POAG), uterine cancer (UtC), abdominal aortic aneurysm (AAA), idiopathic pulmonary fibrosis (IPF), thyroid cancer (ThC).

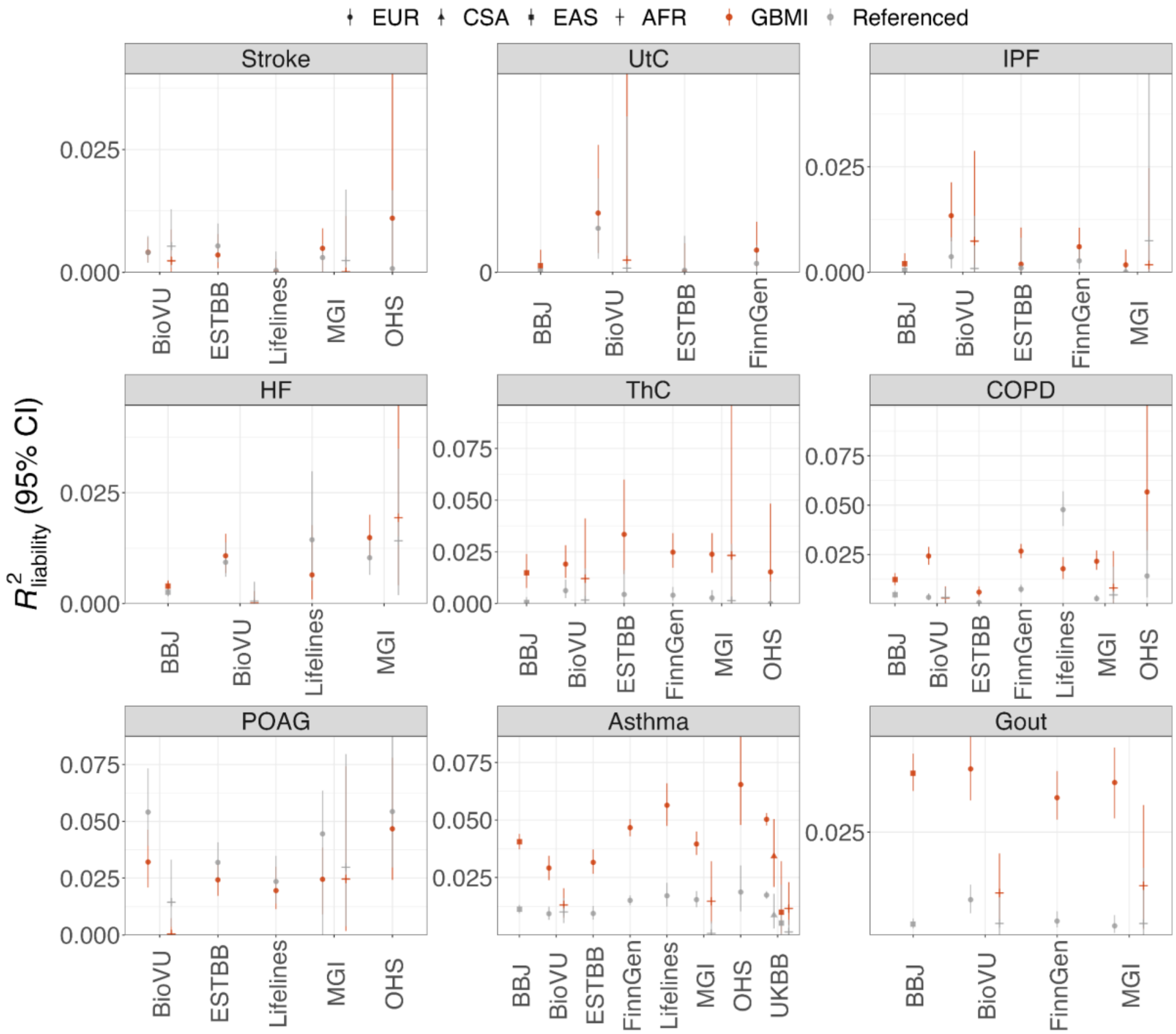

**Figure S12. The prediction performance of GBMI versus previously published GWAS.**

The phenotypes were ranked by the SNP-based heritability estimates from all ancestries. Note that we removed the estimates in AMR and MID due to limited information as a result of small sample sizes. The full results are shown in Table S5. Abbreviations: Europeans (EUR), Central and South Asians (CSA), East Asians (EAS) and Africans (AFR), chronic obstructive pulmonary disease (COPD), heart failure (HF), acute appendicitis (AcApp), venous thromboembolism (VTE), primary open-angle glaucoma (POAG), uterine cancer (UtC), abdominal aortic aneurysm (AAA), idiopathic pulmonary fibrosis (IPF), thyroid cancer (ThC).

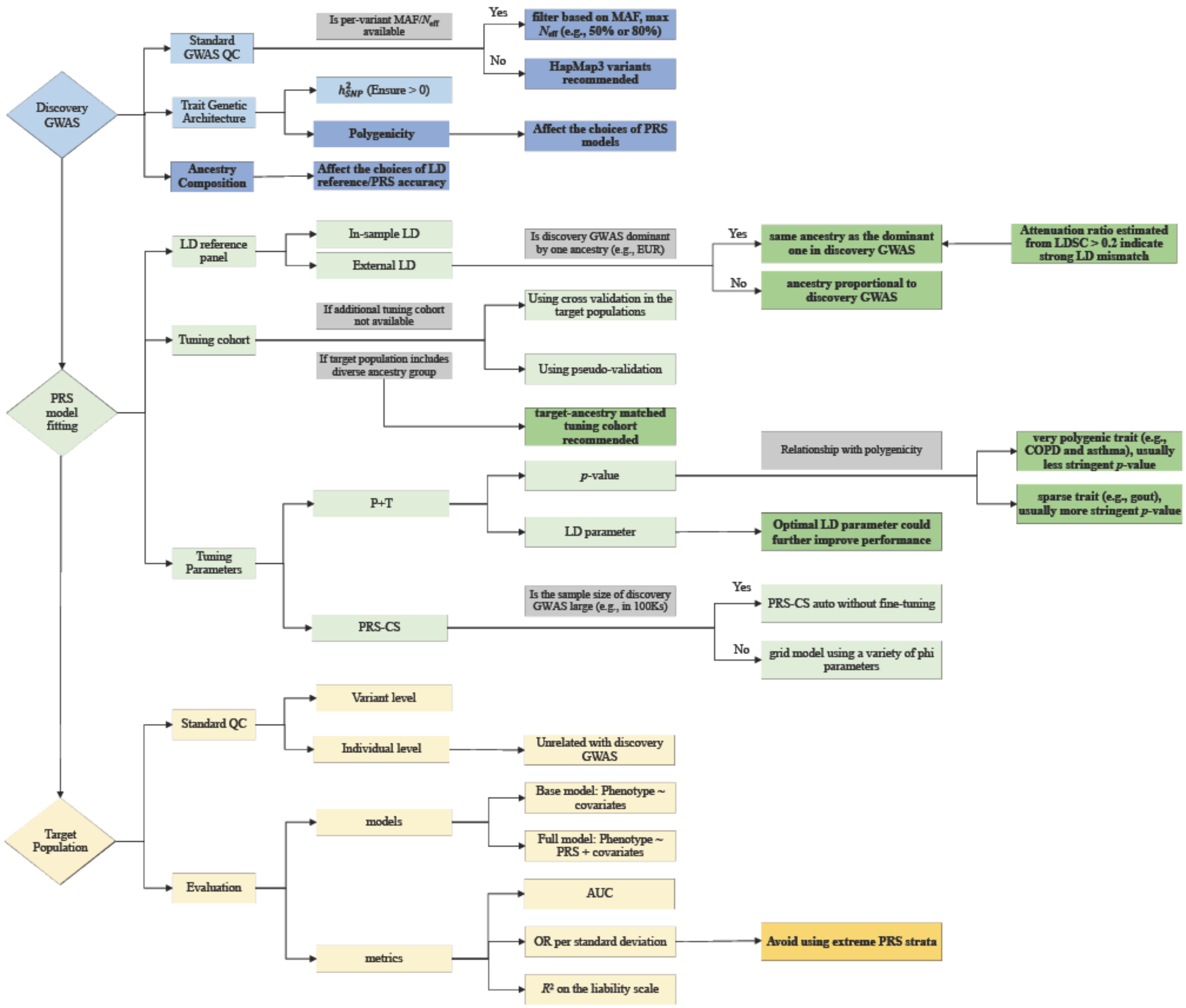

**Figure S13. Flow chart for general lessons and guidelines of best practice using multi-ancestry GWAS for PRS analyses.**

We focused on disease traits where the discovery GWAS is meta-analyzed multi-ancestry GWAS. We used P+T and PRS-CS as examples to show different aspects related to PRS construction. We included three general considerations involved in PRS analyses, including discovery GWAS (blue), PRS model fitting (green) and target populations (yellow). For each of the three general considerations, we highlighted the extended recommendations based on findings in this study in darker color with bold texts. The gray boxes are used for decision making. The detailed study design is shown in **Figure 1**.

### Supplementary Tables

**Table S1. Previously published GWAS used in comparison to GBMI GWAS.**

| Endpoints | Abb. | Public GWAS | Neff | Prevalence | Ancestry Composition |
|---|---|---|---|---|---|
| Asthma | Asthma | [1] | 79722 | 0.168 | EUR: 89.6%, 19965/107715 cases/controls; EAS: 3.7%, 1239/3976 cases/controls; AFR: 5.8%, 2149/6055 cases/controls; AMR: 1.0%, 606/792 cases/controls |
| Chronic obstructive pulmonary disease | COPD | [2] | 75457 | 0.105 | EUR: 100%, 21077/179689 cases/controls |
| Heart Failure | HF | [3] | 180076 | 0.048 | EUR: 100%, 47309/930014 cases/controls |
| Stroke | Stroke | [4] | 233792 | 0.130 | EUR: 86.3%, 40585/406111 cases/controls; CSA: 1.8%, 2437/6707 cases/controls; EAS: 8.8%, 17369/28195 cases/controls; AFR: 4.0%, 541/15154 cases/controls; AMR: 0.3%, 865/692 cases/controls; Mixed Asian: 0.1%, 365/333 cases/controls |
| Acute appendicitis | AcApp | | | | |
| Venous thromboembolism | VTE | | | | |
| Gout | Gout | [5] | 8202 | 0.030 | EUR: 100%, 2115/67259 cases/controls |
| Appendectomy | Appendectomy | | | | |
| Primary open-angle glaucoma | POAG | [6] | 124531 | 0.089 | EUR: 86.3%, 23963/306942 cases/controls; EAS: 12.1%, 6935/39588 cases/controls; AFR: 1.6%, 3281/2791 cases/controls |
| Uterine cancer | UtC | [7] | 8073 | 0.009 | EUR: 100%, 2037/219656 cases/controls |
| Abdominal aortic aneurysm | AAA | | | | |
| Idiopathic pulmonary fibrosis | IPF | [8] | 5459 | 0.003 | EUR: 100%, 1369/435866 cases/controls |
| Thyroid cancer | ThC | [7] | 3042 | 0.002 | EUR: 100%, 762/410350 cases/controls |

**Table S5. Summary of extending practical considerations in PRS analyses in this study.**

| Considerations | Details | Existing Practice (single ancestry) | Extensions (this study) |
|---|---|---|---|
| Discovery GWAS | polygenicity (Proportion of SNPs with non-zero effects) | No consideration of polygenicity regarding PRS model selection | 1) Choose models adaptive to trait genetic architecture; 2) if not, then choose hyper-parameters reflecting polygenicity, such as smaller phi values for less polygenic traits in PRS-CS grid model and larger values for more polygenic traits |
| | SNP-based heritability | Not routinely checked | Should confirm it is significantly larger than 0 |
| | Ancestry composition | Not applicable for single-ancestry GWAS | Informative for the choice of LD reference panel as well as benchmarking PRS accuracy |
| | Per-variant effective sample sizes/MAF | Generally only use MAF filter | additionally apply per-variant effective sample size filter, if such information not available then use HapMap3 variants |
| PRS model fitting | LD reference panel | In-sample LD is preferred, if not available then use ancestry-matched LD reference panel | When using external LD reference: 1) ancestry matched with the dominant one in the discovery GWAS; 2) when no ancestry is dominant, reference panel proportional to discovery GWAS is recommended |
| | Tuning cohort | Use additional tuning cohort if applicable; otherwise using pseudo-validation or splitting target cohort as two parts | When the target population include diverse ancestries, using target-ancestry matched tuning cohort |
| | Tuning parameters | Usually *p*-value threshold for P+T; phi parameter for PRS-CS grid models | Additional LD parameter optimization could slightly improve performance; PRS-CS auto model can be used when discovery GWAS is large enough |
| Target populations | Standard QC | Variant/Individual level; make sure the individuals are unrelated with discovery GWAS | Same, but perform QC per ancestry if diverse ancestries included in the target population |
| | Evaluation | Account for the contribution of PRS by regressing out the effects covariates; reporting the accuracy using different evaluation metrics | Recommend reporting the PRS distribution to benchmark against other predictors; relative accuracy is often reported when diverse ancestries included |

#### References

1. Demenais, F. *et al.* Multiancestry association study identifies new asthma risk loci that colocalize with immune-cell enhancer marks. *Nat. Genet.* **50**, 42–53 (2017).
2. Kim, W. *et al.* Genome-Wide Gene-by-Smoking Interaction Study of Chronic Obstructive Pulmonary Disease. *Am. J. Epidemiol.* **190**, 875–885 (2021).
3. Shah, S. *et al.* Genome-wide association and Mendelian randomisation analysis provide insights into the pathogenesis of heart failure. *Nat. Commun.* **11**, 1–12 (2020).
4. Malik, R. *et al.* Multiancestry genome-wide association study of 520,000 subjects identifies 32 loci associated with stroke and stroke subtypes. *Nat. Genet.* **50**, 524–537 (2018).
5. Köttgen, A. *et al.* Genome-wide association analyses identify 18 new loci associated with serum urate concentrations. *Nat. Genet.* **45**, 145–154 (2013).
6. Gharahkhani, P. *et al.* Genome-wide meta-analysis identifies 127 open-angle glaucoma loci with consistent effect across ancestries. *Nat. Commun.* **12**, 1–16 (2021).
7. Rashkin, S. R. *et al.* Pan-cancer study detects genetic risk variants and shared genetic basis in two large cohorts. *Nat. Commun.* **11**, 1–14 (2020).
8. Duckworth, A. *et al.* A Mendelian randomisation study of smoking causality in IPF compared with COPD. *medRxiv* (2020).
